# Epidemiological and Clinical Characteristics, and Virologic Features of COVID-19 Patients in Kazakhstan: a Nation-Wide, Retrospective, Cohort Study

**DOI:** 10.1101/2021.01.06.20249091

**Authors:** Sergey Yegorov, Maiya Goremykina, Raifa Ivanova, Sara V. Good, Dmitriy Babenko, Alexandr Shevtsov, on behalf of the COVID-19 Genomics Research Group, Kelly S. MacDonald, Yersin Zhunussov, on behalf of the Semey COVID-19 Epidemiology Research Group

## Abstract

**Background:** The earliest coronavirus disease-2019 (COVID-19) cases in Central Asia were announced in March 2020 by Kazakhstan. Despite the implementation of aggressive measures to curb infection spread, gaps remain in the understanding of the clinical and epidemiologic features of the regional pandemic.

**Methods:** We did a retrospective, observational cohort study of patients with laboratory-confirmed COVID-19 in Kazakhstan between February and April 2020. We compared demographic, clinical, laboratory and radiological data of patients with different COVID-19 severities on admission. Univariable and multivariable logistic regression was used to assess factors associated with disease severity and death. Whole-genome SARS-CoV-2 analysis was performed in 53 patients without a recent history of international travel.

**Findings:** Of the 1072 patients with laboratory-confirmed COVID-19 in March-April 2020, the median age was 36 years (IQR 24–50) and 484 (45%) were male. On admission, 683 (64%) participants had mild, 341 (32%) moderate, and 47 (4%) severe-to-critical COVID-19 manifestation; 20 deaths (1.87%) were reported at study exit. Multivariable regression indicated increasing odds of severe disease associated with older age (odds ratio 1.05, 95% CI 1.03-1.07, per year increase; p<0.001), the presence of comorbidities (2.13, 95% CI 1.07-4.23; p<0.031) and elevated white blood cell count (WBC, 1.14, 95% CI 1.01-1.28; p<0.032) on admission, while older age (1.09, 95% CI 1.06-1.12, per year increase; p<0.001) and male sex (5.97, 95% CI 1.95-18.32; p<0.002) were associated with increased odds of death. The Kazakhstan SARS-CoV-2 isolates grouped into seven distinct lineages O/B.4.1, S/A.2, S/B.1.1, G/B.1, GH/B.1.255, GH/B.1.3 and GR/B.1.1.10.

**Interpretation:** Older age, comorbidities, increased WBC count, and male sex were risk factors for COVID-19 disease severity and mortality in Kazakhstan. The broad SARS-CoV-2 diversity suggests multiple importations and community-level amplification, likely predating the declaration of state emergency. Continuous epidemiologic and genomic surveillance may be critical for a better understanding of the regional COVID-19 dynamics.

## INTRODUCTION

Severe Acute Respiratory Syndrome Coronavirus 2 (SARS-CoV-2), the cause of the coronavirus disease-2019 (COVID-19), within months of emergence from Wuhan, China, rapidly spread exacting a devastating human toll across the globe ^1^. While the search for effective treatments continues and vaccines have commenced early implementation ^1^, it is imperative that up-to-date information be available from diverse populations on the disease epidemiology, clinical presentation, and population-specific characteristics influencing COVID-19 prevention, treatment, and vaccine strategies. This is especially critical in low- and middle-income countries (LMIC), where epidemiologic surveillance is often constrained due to resource shortages ^2^.

Kazakhstan was the first among the Central Asian LMICs to initiate COVID-19 screening in early 2020, with the first confirmed cases identified in mid-March, and a country-wide emergency state declared on 16 March 2020 ^3^. Kazakhstan neighbours Russia, China, and the Central Asian states, and harbours an extensive ground and airway transit network, setting it in a vulnerable position for both COVID-19 importation and rapid community spread. Despite the initial swift public health response, Kazakhstan has encountered major barriers regarding case reporting and attempts have been made by both the government and public to increase transparency over COVID-19-related morbidity and mortality ^3,4^. Thus a recent study from Kazakhstan has described the clinical characteristics of COVID-19 in children ^5^, corroborating a relatively mild disease course for pediatric COVID-19.

However, little is still known about the clinical attributes and molecular epidemiology of COVID-19 across varied age and ethnic groups, information that is urgently needed to guide the public health authorities and clinicians in the wake of the on-going pandemic. Here, we examined data from hospitalized patients with laboratory-confirmed COVID-19 during the first months of the pandemic to explore demographic, clinical and laboratory features and factors associated with COVID-19 disease severity and death in Kazakhstan. We also analyzed whole-genome SARS-CoV-2 data to characterize the regional community-level virus diversity.

## METHODS

### Study design and participants

Medical records were obtained for individuals who presented with COVID-19-like symptoms or had a suspected exposure to SARS-CoV-2 between 20 February and 30 April 2020, as reported to the Republican Centre for Health Development by hospitals in 14 regions and 3 major cities (Fig 1A). During this period, all subjects with suspected or confirmed COVID-19 infection were hospitalized in specialized “provisional” clinics. The retrospective cohort study was approved by the Research Ethics Board of Semey Medical University as an anonymized epidemiological study, for which the requirement for informed consent was waived due to the pandemic state and urgent need to collect and analyze data. Virological samples used in genomic studies were anonymized and all genomic study procedures were approved by the Ethics Committee of the National Centre for Biotechnology.

**Figure 1:**
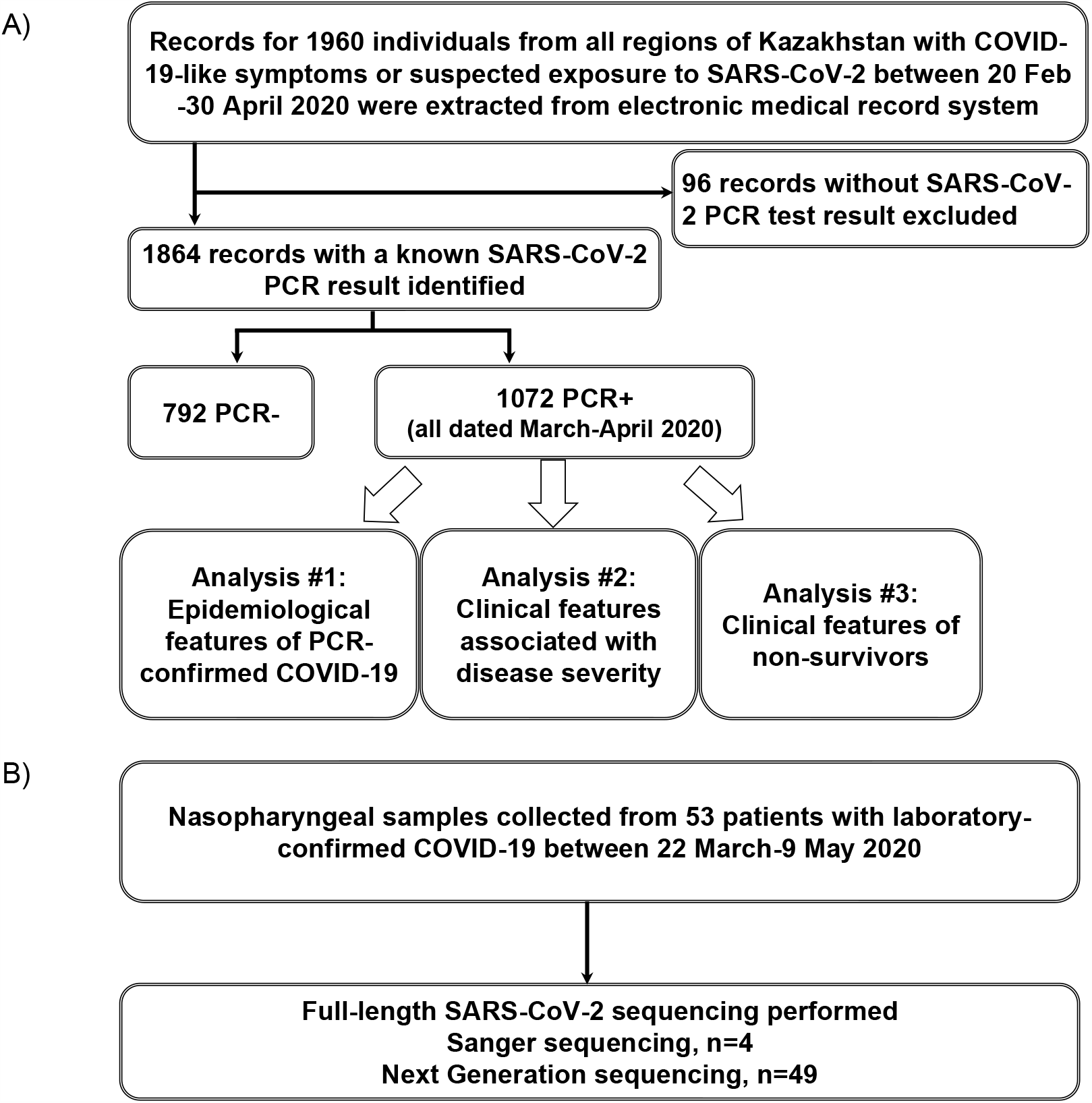
**A**. The Retrospective cohort study profile, **B**. The SARS-CoV-2 genomic study profile.

**Figure 2:**
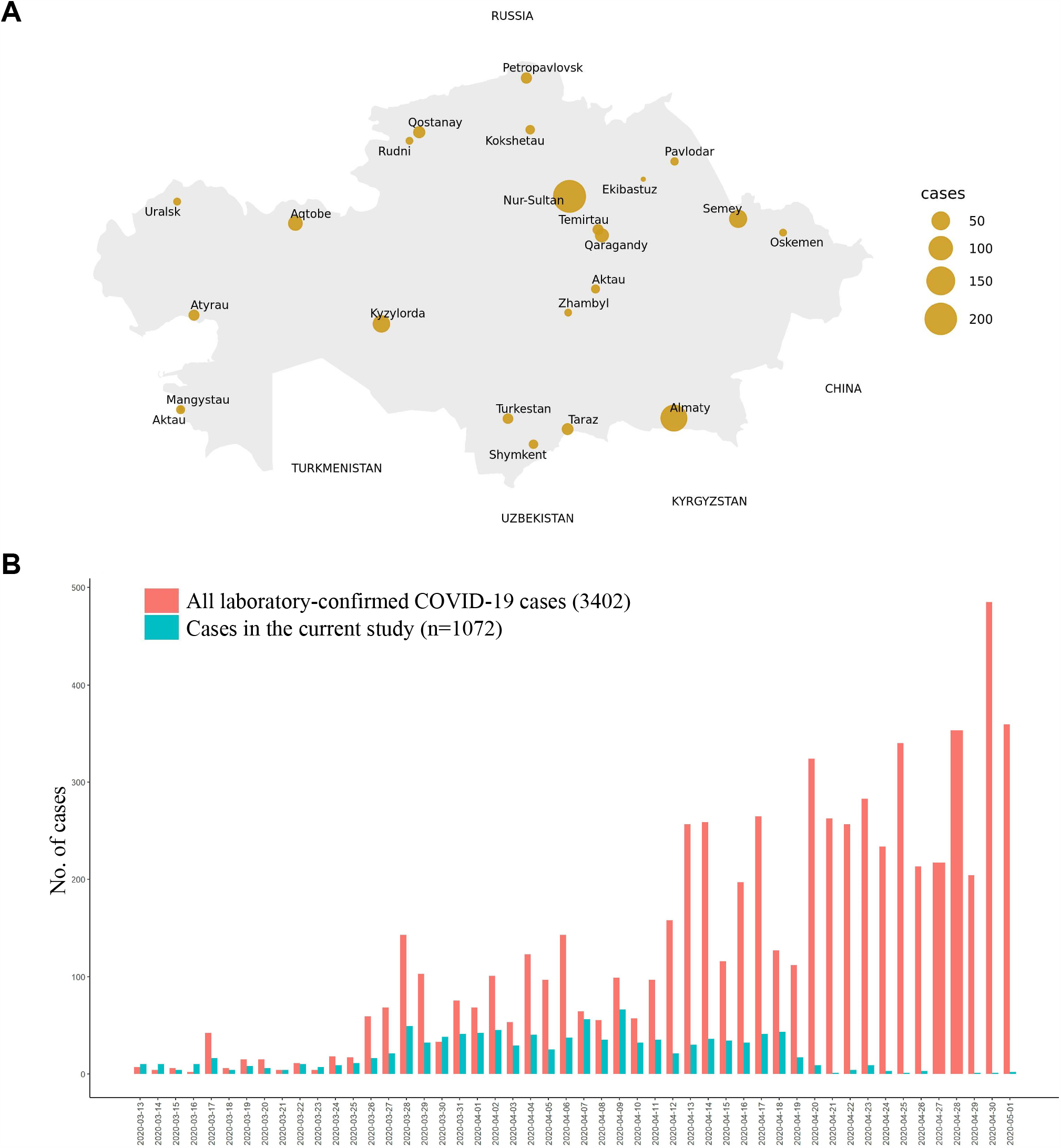
**A**. Distribution of patients with laboratory-confirmed COVID-19 across Kazakhstan. Based on data for 574 patients, for whom the site of initial diagnosis was known in the current study. **B**. Epidemic curve of the confirmed COVID-19 cases in the current study compared to the official statistics on confirmed COVID-19 cases in Kazakhstan in March-April 2020 according to the Republican Centre for Health Development and World Health Organization (WHO). Official statistics were obtained from the WHO website ^14^.

### Data collection

Epidemiological, demographic, clinical, laboratory, treatment, and outcome data were extracted from de-identified electronic medical records using a standardised data collection form. Radiological data were only available in the form of a radiologist’s final diagnosis; a detailed description of imaging scans was unavailable. All data were entered into a computerized database and cross-checked by two physicians (MG and RI), and subsequently by a third researcher (SY).

### Laboratory procedures

Laboratory confirmation of SARS-CoV-2 infection was done using real-time quantitative - PCR (qPCR) on nasopharyngeal swabs by the regional National Centre of Expertise (NCE) laboratories using the Beijing Genomics Institute (BGI) kit (Shenzhen, China) targeting the Orf1ab locus A. Laboratory examinations included complete blood counts, blood chemical analyses, coagulation testing, liver and renal function assessment, and measurements of electrolytes, C-reactive protein, creatinine, D-dimer and lactate.

### Virus genome sequencing and bioinformatics

Nasopharyngeal swabs were collected from 53 randomly selected symptomatic patients, who had a positive SARS-CoV-2 PCR test and were hospitalized in the capital city, Nur-Sultan between 22 March and 9 May (Fig 1B). Travel history was available for 49 patients, none of whom had a history of recent international travel. Full description of the sequencing methods involving Sanger and Next Generation Illumina Sequencing is provided in appendix pp. 2-4.

### Phylogenetic analyses

The global SARS-CoV2 phylogeny, consisting of 5022 genomes representative of the total repository of 66,940 sequences, and associated metadata were downloaded from Nextstrain^6^ on 8 July 2020; the tree was reconstructed using the Nextstrain/ncov pipeline. To visualize the phylogenetic relationship of the Kazakhstan isolates in the context of possible origins of importation, we created a tree using 165 sequences from the Global Initiative on Sharing All Influenza Data (GISAID) ^7^ satisfying one or all of the following criteria: i) proximity to the Kazakhstan sequences, as determined by calculating the pairwise distance between all samples in the Global GISAID Phylogeny using snp-dists ^8^ and selecting the global strains closest to the Kazakhstan isolates (n=21); ii) representing non-redundant SARS-CoV-2 strains from regions, where international travel was reported from in the current cohort (see Fig 3) (n=63); and iii) randomly selected SARS-CoV-2 full-length sequences from each of the clades observed in Kazakhstan (appendix p. 5) per month between 31 December 2019 and 9 May 2020 (n=81). Sequences were aligned to the Wuhan reference using MAFFT ^9^, the alignment checked for discrepancies and ends trimmed manually to match the reference, followed by reconstruction of a maximum likelihood phylogeny in IQTREE ^10^ using the GTR + F + I model and ultrafast bootstrap option with 1,000 replicates. The phylogenetic trees were visualized using ggtree ^11^, and sequences classified using the GISAID and dynamic nomenclature systems ^12^.

**Figure 3:**
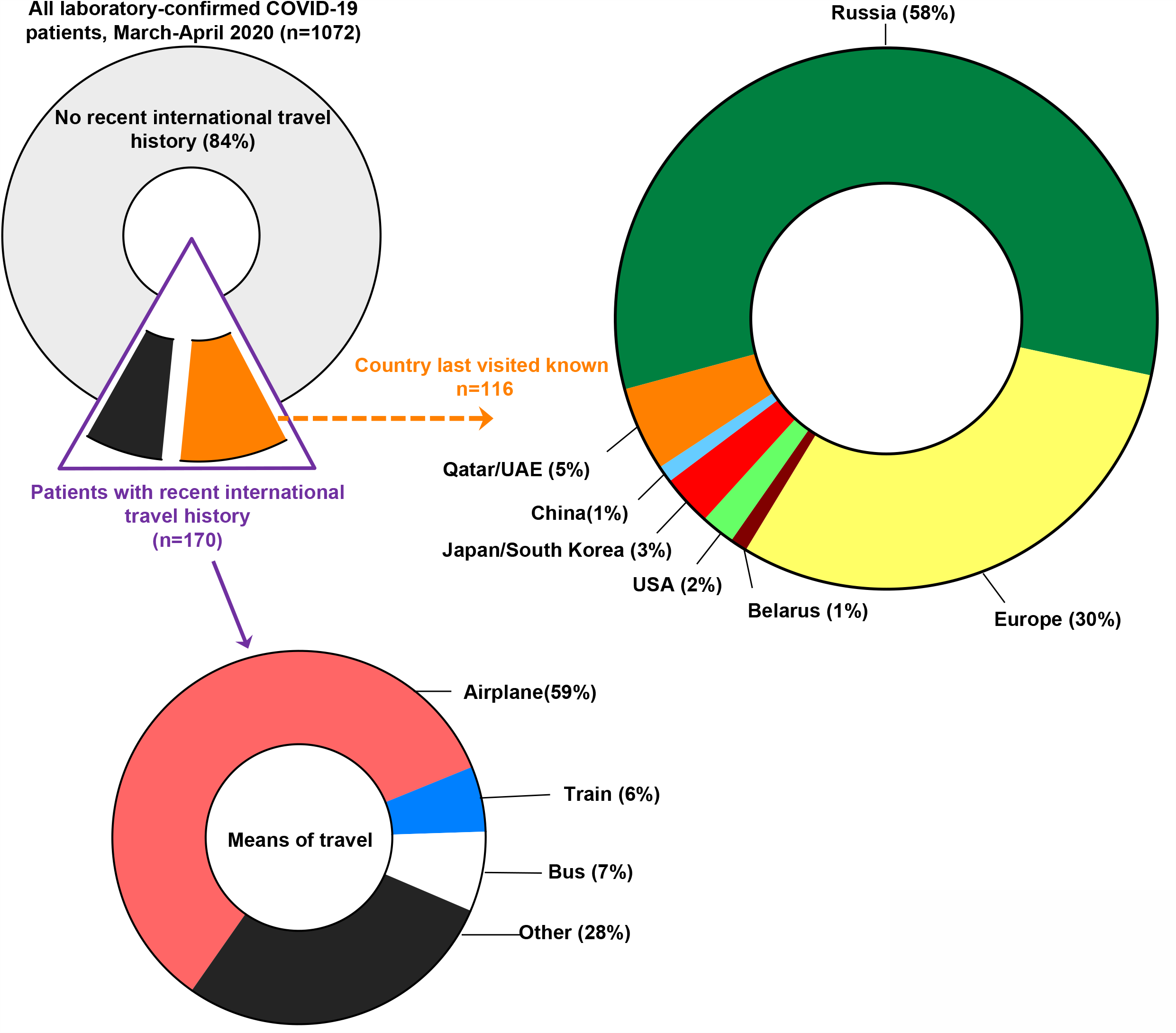
Regions of travel and transportation used by laboratory-confirmed COVID-19 patients with a recent history of international travel.

### Definitions

A patient was assigned a laboratory-confirmed COVID-19 diagnosis if their medical record contained at least one positive SARS-CoV-2 PCR test result. Fever was defined as axillary temperature of at least 37□3°C. The degree of COVID-19 severity was defined following the interim WHO guidelines ^13^. To avoid inconsistencies arising from varied terminology in electronic records, patients described as having “asymptomatic”, “presymptomatic” and “mild” COVID-19 severity were combined into one “mild COVID-19 disease” category. To facilitate comparisons with the literature, the term “non-severe” COVID-19 was used for both “mild” and “moderate” disease, while the term “severe” COVID-19 was reserved for the pooled “severe” and “critical” groups. Recent international travel history was defined as travel outside of Kazakhstan within two weeks prior to hospitalization or symptom onset, whichever was first.

### Statistical analysis

Variables were excluded prior to analysis if data were available for less than 40% of patients in any comparison group. We used the two-sided Mann-Whitney U, χ2, or Fisher’s exact tests to compare differences between groups, as appropriate. For uni- and multivariable logistic regression analyses, patients with mild and moderate disease severity were pooled into one “non-severe” category and compared with the “severe” (severe-critical) patient group. Risk factors associated with disease severity and death and their odds ratios (OR) were first analyzed by univariable logistic regression. In the multivariable logistic regression analyses, to avoid model overfitting due to the limited size of endpoint events, four variables were chosen for the analysis of disease severity (n=47 for severe-critical patients) and two variables were ultimately included in the non-survivor analysis (n=20 for non-survivors). Variables for multivariable analyses were selected based on significance in the univariable model (p<0.05), low collinearity, sample size constraints for each variable, and prior knowledge from the literature. Statistical analyses were performed using IBM SPSS V.23 and R V.3.6.3.

## RESULTS

Medical record data were obtained for a total of 1960 subjects with COVID-19-like clinical symptoms or suspected exposure to SARS-CoV-2 via a close contact or travel. After excluding 96 patients, for whom a SARS-CoV-2 PCR result was unavailable, there were 1864 patients with a known PCR result (Fig 1A). Our analysis focused on the 1072 patients, who had a confirmed SARS-CoV-2 PCR+ diagnosis, representing 32% of all (n=3402) patients with laboratory confirmed COVID-19 (Fig 2A), who had been hospitalized in Kazakhstan as of April 30, 2020 ^14^.

The largest number of PCR-confirmed patients (207, 19%) were admitted in the capital city and affiliated municipality (Nur-Sultan, Akmola region), followed by Almaty city and region (127, 12%) (Fig 2B). 344 (32%) and 728 (68%) patients were hospitalized in March and April, respectively, and all patients had been discharged or died by 5 May 2020. 439 (41%) of all PCR+ cases were identified through COVID-19 screening of inbound travelers and through tracing activities, while 633 (59%) patients were admitted based on the presence of COVID-19-like symptomatology. Professional occupation data were available for 350 patients, of whom 91 (26%) were identified as health care workers. Recent international travel was reported by 170 (16%) patients; most reported travel was from Russia (58%) and Europe (30%), via airplane (59%) and ground transportation (41%) (Fig 3). The comparisons of patient demographic and clinical characteristics on admission grouped by disease severity and death are shown in Tables 1 and 2, respectively.

**Table 1:** Demographic and clinical characteristics of laboratory-confirmed COVID-19 patients categorized on admission by disease severity.

|  | **All (1072)** | **Mild (683)** | **Moderate (341)** | **Severe/critical (47)** | **P value** |
| --- | --- | --- | --- | --- | --- |
| Age, years | 36.0 (24.0-50.0) | 32.0 (23.0-47.0) | 38.0 (28.0-53.0) | 58.0 (45.5-71.0) | <0.001 |
| 0-14 | 94 (8.77%) | 76 (11.1%) | 17 (4.99%) | 1 (2.13%) | <0.001 |
| 15-49 | 695 (64.8%) | 458 (67.1%) | 222 (65.1%) | 15 (31.9%) |  |
| 50-64 | 202 (18.8%) | 113 (16.5%) | 76 (22.3%) | 13 (27.7%) |  |
| ≥ 65 | 81 (7.56%) | 36 (5.27%) | 26 (7.62%) | 18 (38.3%) |  |
| Sex (Men) | 484 (45.1%) | 301 (44.1%) | 155 (45.5%) | 27 (57.4%) | 0.157 |
| Kazakh ethnicity | 852 (79.5%) | 551 (80.7%) | 271 (79.5%) | 30 (62.5%) | 0.011 |
| BMI* | 23.9 (21.4-28.7) | 23.0 (20.0-26.2) | 25.8 (23.1-30.7) | 27.5 (25.8-29.2) | 0.012 |
| Days from symptom onset to admission | 1 (1-3) | 1 (1-2) | 2 (1-6) | 4 (2-7) | <0.001 |
| Days in Hospital | 16.0 (14.0-17.0) | 16.0 (14.0-17.0) | 16.0 (15.0-18.0) | 15.0 (3.00-17.0) | 0.075 |
| Deaths | 20 (1.9%) | 1 (0.1%) | 4 (1.2%) | 15 (31.3%) | <0.001 |
| **Comorbidities (any)** | 431 (40.2%) | 235 (34.4%) | 163 (47.8%) | 33 (68.8%) | <0.001 |
| *Hypertension* | 98 (9.14%) | 37 (5.42%) | 44 (12.9%) | 17 (36.2%) | <0.001 |
| *Coronary heart disease* | 20 (1.87%) | 8 (1.17%) | 10 (2.93%) | 2 (4.26%) | 0.07 |
| *COPD* | 6 (0.56%) | 2 (0.29%) | 2 (0.59%) | 2 (4.26%) | 0.002 |
| *Chronic kidney disease* | 29 (2.71%) | 11 (1.61%) | 15 (4.40%) | 3 (6.38%) | 0.01 |
| *Cancer* | 6 (0.56%) | 2 (0.29%) | 3 (0.88%) | 1 (2.13%) | 0.173 |
| *Diabetes* | 26 (2.43%) | 9 (1.32%) | 9 (2.64%) | 8 (17.0%) | <0.001 |
| *Liver disease* | 15 (1.40%) | 2 (0.29%) | 7 (2.05%) | 6 (12.8%) | <0.001 |
| *Other comorbidity* | 364<br>(34.0%) | 200 (29.3%) | 135<br>(39.6%) | 29 (61.7%) | <0.001 |
| **Signs and symptoms** |  |  |  |  |  |
| *Median body temperature\** | 36.6 (36.4-37.0) | 36.5 (36.3-36.7) | 36.8 (36.5-37.4) | 37.8 (36.8-38.0) | <0.001 |
| *Fever (ax temp ≥ 37.3C)* | 131<br>(12.2%) | 31 (4.54%) | 82 (24.0%) | 18 (38.3%) | <0.001 |
| *Cough* | 294<br>(27.4%) | 104 (15.2%) | 163<br>(47.8%) | 27 (57.4%) | <0.001 |
| *Sputum production* | 59 (5.50%) | 20 (2.93%) | 30 (8.80%) | 9 (19.1%) | <0.001 |
| *Shortness of breath* | 56 (5.22%) | 6 (0.88%) | 30 (8.80%) | 20 (42.6%) | <0.001 |
| *Dyspnoea* | 31 (2.89%) | 3 (0.44%) | 17 (4.99%) | 11 (23.4%) | <0.001 |
| *Stuffy/runny nose* | 90 (8.40%) | 45 (6.59%) | 40 (11.7%) | 5 (10.6%) | 0.018 |
| *Sore throat* | 128<br>(11.9%) | 53 (7.76%) | 61 (17.9%) | 14 (29.8%) | <0.001 |
| *Oropharynx hyperemia* | 391<br>(36.5%) | 221 (32.4%) | 149<br>(43.7%) | 21 (44.7%) | 0.001 |
| *Tonsill hypertrophy* | 63 (5.88%) | 24 (3.51%) | 32 (9.38%) | 7 (14.9%) | <0.001 |
| *Chest pain* | 46 (4.29%) | 10 (1.46%) | 27 (7.92%) | 9 (19.1%) | <0.001 |
| *Chest tightness* | 21 (1.96%) | 5 (0.73%) | 11 (3.23%) | 5 (10.6%) | <0.001 |
| *Wheezing* | 44 (4.10%) | 11 (1.61%) | 18 (5.28%) | 15 (31.9%) | <0.001 |
| *Diarrhoea* | 27 (2.52%) | 10 (1.46%) | 15 (4.40%) | 2 (4.26%) | 0.014 |
| *Nausea/vomiting* | 23 (2.15%) | 5 (0.73%) | 13 (3.81%) | 5 (10.6%) | <0.001 |
| *Headache* | 113<br>(10.5%) | 35 (5.12%) | 64 (18.8%) | 14 (29.8%) | <0.001 |
| *Conjunctivitis* | 5 (0.47%) | 2 (0.29%) | 2 (0.59%) | 1 (2.13%) | 0.197 |
| *Myalgia/fatigue* | 433<br>(40.4%) | 251 (36.7%) | 155<br>(45.5%) | 27 (57.4%) | 0.002 |
| *Joint pain* | 21 (1·96%) | 8 (1·17%) | 10 (2·93%) | 3 (6·38%) | 0·014 |
| *Median pulse\*~* | 80·0 (76·0-87·0) | 80·0 (74·0-85·0) | 81·0 (78·0-88·0) | 89·0 (80·0-103) | 0·057 |
| *>125 beats/min* | 8 (0·75%) | 3 (0·44%) | 3 (0·88%) | 2 (4·26%) | <0·001 |
| *Systolic pressure \** | 110 (110-120) | 110 (110-120) | 120 (110-130) | 118 (108-126) | <0·001 |
| *<90 mmHg* | 11 (1·03%) | 8 (1·17%) | 0 (0·00%) | 3 (6·38%) | <0·001 |
| *Diastolic pressure \** | 76·0 (70·0-80·0) | 70·0 (70·0-80·0) | 80·0 (70·0-80·0) | 70·0 (60·0-80·0) | <0·001 |
| *Respiratory rate \** | 18·0 (18·0-20·0) | 18·0 (18·0-19·0) | 18·0 (18·0-20·0) | 22·5 (20·0-28·5) | <0·001 |
| *>24 breaths/min* | 25 (2·33%) | 8 (1·17%) | 8 (2·35%) | 9 (19·1%) | <0·001 |
| *SpO2 \** | 97·0 (96·0-98·0) | 98·0 (96·0-98·0) | 97·0 (96·0-98·0) | 91·5 (84·5-95·0) | <0·001 |
| **Laboratory findings** |  |  |  |  |  |
| White blood cells, x10<sup>9</sup>/L, median | 6·30 (5·10-7·90) | 6·47 (5·10-8·00) | 6·10 (5·00-7·68) | 6·55 (5·50-9·05) | 0·067 |
| *< 4 x10<sup>9</sup>/L* | 90 (8·8%) | 54 (8·3%) | 34 (10·4%) | 2 (4·4%) | 0·056 |
| *> 10 x10<sup>9</sup>/L* | 79 (7·8%) | 51 (7·9%) | 20 (6·1%) | 8 (17·8%) |  |
| Neutrophil count, x10<sup>9</sup>/L, median * | 3·80 (2·67-5·49) | 3·84 (2·74-5·40) | 3·66 (2·68-5·43) | 5·14 (2·29-10·6) | 0·404 |
| *< 2 x10<sup>9</sup>/L* | 64 (13·3%) | 35 (11·8%) | 23 (14·6%) | 6 (20·7%) | 0·33 |
| Lymphocyte count, x10<sup>9</sup>/L, median * | 1·73 (1·09-2·54) | 1·76 (1·13-2·52) | 1·68 (1·03-2·50) | 1·75 (1·00-2·91) | 0·523 |
| *< 1 x10<sup>9</sup>/L* | 160 (23·3%) | 93 (22·4%) | 58 (24·7%) | 9 (25·7%) | 0·751 |
| NLR * | 2·06 (1·36-3·16) | 2·00 (1·38-3·14) | 2·10 (1·45-3·05) | 2·58 (0·91-6·48) | 0·581 |
| *> 4·0* | 76 (16·3%) | 45 (15·7%) | 22 (14·4%) | 9 (33·3%) | 0·044 |
| Median haemoglobin, g/L | 136 (124-150) | 137 (124-149) | 137 (124-152) | 127 (114-142) | 0·007 |
| Monocytes, x10<sup>9</sup>/L, median * | 0.38 (0.17-0.70) | 0.40 (0.17-0.68) | 0.33 (0.17-0.70) | 0.41 (0.25-1.28) | 0.384 |
| Eosinophils, x10<sup>9</sup>/L, median * | 2.00 (1.00-3.00) | 1.00 (0.61-3.00) | 2.00 (1.00-3.00) | 1.50 (0.88-4.00) | 0.145 |
| Platelet count, x10<sup>9</sup>/L, median * | 236 (196-288) | 240 (200-295) | 230 (190-275) | 236 (190-304) | 0.099 |
| < 100/L | 8 (0.8%) | 4 (0.7%) | 3 (1.0%) | 1 (2.4%) | 0.465 |
| Median prothrombin time, s * | 13.0 (11.7-16.0) | 12.8 (11.4-16) | 13 (11.85-15.75) | 14.75 (12.48-19.6) | 0.018 |
| Median fibrinogen, g/L * | 3.10 (2.40-4.50) | 3.1 (2.37-4.2) | 3.04 (2.38-4.38) | 5 (2.93-6.35) | 0.01 |
| Median albumin, g/L* | 40.0 (35.4-44.0) | 40 (37-44.31) | 40 (34.95-44) | 35.2 (24-38.14) | 0.02 |
| Median alanine aminotransferase, U/L * | 18.0 (9.78-33.0) | 16.9 (9.2-32) | 20.8 (11.05-37) | 22.8 (11.5-47) | 0.02 |
| Median aspartate aminotransferase, U/L * | 18.5 (12.5-26.5) | 17.71 (12-23.2) | 20.04 (13.78-29.55) | 38.69 (17.55-50.28) | <0.001 |
| Total bilirubin, mmol/L, median * | 11.2 (8.00-16.0) | 11.5 (8.2-16.73) | 10.45 (7.58-15) | 13.2 (10-20.1) | 0.002 |
| Direct bilirubin, mmol/L, median * | 2.70 (1.79-4.30) | 2.50 (1.72-4.19) | 2.64 (1.70-4.30) | 4.20 (3.45-10.4) | 0.002 |
| Median glucose, mmol/L * | 5.10 (4.50-6.00) | 4.91 (4.4-5.8) | 5.3 (4.6-6.12) | 6.7 (5.02-8.98) | <0.001 |
| Median blood urea nitrogen, mmol/L * | 4.40 (3.47-5.54) | 4.3 (3.4-5.36) | 4.4 (3.56-5.54) | 6.95 (4.27-10.1) | <0.001 |
| Median creatinine, uM* | 73.1 (60.3-88.0) | 70.1 (57.08-86.2) | 75.5 (62.45-92.8) | 84 (67-117) | <0.001 |
| Median C-reactive protein, mg/L * | 2.30 (0.09-8.10) | 2.00 (0.00-6.00) | 2.69 (0.12-10.0) | 6.00 (0.65-45.1) | 0.022 |
| Median sodium, mmol/L* | 141 (138-143) | 141 (139-143) | 140 (137-144) | 139 (135-142) | 0.239 |
| Median potassium, mmol/L * | 4.10 (3.69-4.40) | 4.10 (3.61-4.39) | 4.20 (3.88-4.52) | 3.70 (3.48-4.13) | 0.033 |
| Median calcium, mmol/L | 2.20 (1.20-2.38) | 2.26 (2.17-2.40) | 2.10 (1.17-2.27) | 1.14 (1.07-1.18) | <0.001 |
| **Radiologic findings \*** |  |  |  |  | <0.001 |
| Pneumonia | 230 (31.2%) | 69 (15.6%) | 130 (49.8%) | 31 (91.2%) |  |
| Bronchitis | 188 (25.5%) | 113 (25.6%) | 73 (28.0%) | 2 (5.88%) |  |
| **Treatments** |  |  |  |  |  |
| Antiviral medications | 749 (69.9%) | 461 (67.5%) | 261 (76.5%) | 27 (57.4%) | 0.001 |
| *Ribavirin* | 75 (7.00%) | 15 (2.20%) | 52 (15.2%) | 8 (17.0%) | <0.001 |
| *Lopinavir and ritonavir* | 696 (64.9%) | 434 (63.5%) | 238 (69.8%) | 24 (51.1%) | 0.012 |
| *Oseltamivir* | 30 (2.80%) | 5 (0.73%) | 21 (6.16%) | 4 (8.51%) | <0.001 |
| *Other* | 18 (1.7%) | 7 (1%) | 11 (3.2%) | 0 (0) | 0.023 |
| Antibiotics | 308 (28.7%) | 114 (16.7%) | 168 (49.3%) | 26 (55.3%) | <0.001 |
| Hydroxychloroquine | 23 (2.15%) | 13 (1.90%) | 9 (2.64%) | 1 (2.13%) | 0.745 |
| Anticoagulant/antiplatelet therapy | 29 (2.71%) | 6 (0.88%) | 12 (3.52%) | 11 (23.4%) | <0.001 |
| Corticosteroids | 16 (1.49%) | 7 (1.02%) | 7 (2.05%) | 2 (4.26%) | 0.13 |
| Oxygen therapy | 64 (6%) | 10 (1.5%) | 23 (6.7) | 31 (64.6%) | <0.001 |
| Mechanical ventilation | 27 (2.5%) | 2 (0.3%) | 6 (1.8%) | 19 (39.6%) | <0.001 |

Data are median (IQR) or n/N (%), where N is the number of patients with available data.
BMI=body mass index. COPD= Chronic obstructive pulmonary disease.
NLR=neutrophil/lymphocyte ratio. SpO2=oxygen saturation. \*Data were available for <1072 patients, see appendix p. 6 for details on available sample size.

**Table 2:** Demographic and clinical characteristics of laboratory-confirmed COVID-19 patients, who had survived (survivors) or died (non-survivors) by 30 April 2020.

|  | All (1072) | Survivors (1052) | Non-survivors | p value |
| --- | --- | --- | --- | --- |

|  |  |  | (N=20) |  |
| --- | --- | --- | --- | --- |
| Age, years | 36.0 (24.0-50.0) | 35.0 (24.0-50.0) | 65.0 (57.8-77.8) | <0.001 |
| 0-14 | 94 (8.77%) | 93 (8.84%) | 1 (5.00%) |  |
| 15-49 | 695 (64.8%) | 693 (65.9%) | 2 (10.0%) |  |
| 50-64 | 202 (18.8%) | 194 (18.4%) | 8 (40.0%) |  |
| ≥ 65 | 81 (7.56%) | 72 (6.84%) | 9 (45.0%) |  |
| Sex (Men) | 484 (45.1%) | 470 (44.7%) | 14 (70.0%) | 0.043 |
| Kazakh ethnicity | 852 (79.5%) | 842 (80.0%) | 10 (50.0%) | 0.001 |
| BMI* | 23.9 (21.4-28.7) | 23.9 (21.4-28.7) | 43.5 (33.8-53.1) | 0.213 |
| Days from symptom onset to admission | 1 (1-3) | 1 (1-3) | 4 (3-10) | <0.001 |
| Days in Hospital | 16.0 (14.0-17.0) | 16.0 (14.0-17.0) | 4 (2.0-15.0) | <0.001 |
| **Comorbidities** | 431 (40.2%) | 415 (39.4%) | 16 (80.0%) | <0.001 |
| *Hypertension* | 98 (9.14%) | 87 (8.27%) | 11 (55.0%) | <0.001 |
| *Coronary heart disease* | 20 (1.87%) | 18 (1.71%) | 2 (10.0%) | 0.051 |
| *COPD* | 6 (0.56%) | 5 (0.48%) | 1 (5.00%) | 0.107 |
| *Chronic kidney disease* | 29 (2.71%) | 26 (2.47%) | 3 (15.0%) | 0.015 |
| *Cancer* | 6 (0.56%) | 6 (0.57%) | 0 (0.00%) | 1.000 |
| *Diabetes* | 26 (2.43%) | 23 (2.19%) | 3 (15.0%) | 0.011 |
| *Liver disease* | 15 (1.40%) | 11 (1.05%) | 4 (20.0%) | <0.001 |
| *Other comorbidity* | 364 (34.0%) | 350 (33.3%) | 14 (70.0%) | 0.001 |
| **Signs and symptoms** |  |  |  |  |
| *Median body temperature\** | 36.6 (36.4-37.0) | 36.6 (36.4-36.9) | 38.0 (37.8-38.2) | 0.004 |
| *Fever (ax temp ≥ 37.3C)* | 131 (12.2%) | 124 (11.8%) | 7 (35.0%) | <0.001 |
| *Cough* | 294 (27.4%) | 286 (27.2%) | 8 (40.0%) | 0.308 |
| *Sputum production* | 59 (5.50%) | 56 (5.32%) | 3 (15.0%) | 0.093 |
| *Shortness of breath* | 56 (5.22%) | 49 (4.66%) | 7 (35.0%) | <0.001 |
| *Dyspnoea* | 31 (2.89%) | 25 (2.38%) | 6 (30.0%) | <0.001 |
| *Stuffy/runny nose* | 90 (8.40%) | 87 (8.27%) | 3 (15.0%) | 0.232 |
| *Sore throat* | 128 (11.9%) | 126 (12.0%) | 2 (10.0%) | 1.000 |
| *Oropharynx hyperemia* | 391 (36.5%) | 386 (36.7%) | 5 (25.0%) | 0.400 |
| *Tonsill hypertrophy* | 63 (5.88%) | 62 (5.89%) | 1 (5.00%) | 1.000 |
| *Chest pain* | 46 (4.29%) | 41 (3.90%) | 5 (25.0%) | 0.001 |
| *Chest tightness* | 21 (1.96%) | 18 (1.71%) | 3 (15.0%) | 0.006 |
| *Wheezing* | 44 (4.10%) | 39 (3.71%) | 5 (25.0%) | 0.001 |
| *Diarrhoea* | 27 (2.52%) | 26 (2.47%) | 1 (5.00%) | 0.402 |
| *Nausea/vomiting* | 23 (2.15%) | 22 (2.09%) | 1 (5.00%) | 0.354 |
| *Headache* | 113 (10.5%) | 108 (10.3%) | 5 (25.0%) | 0.051 |
| *Conjunctivitis* | 5 (0.47%) | 4 (0.38%) | 1 (5.00%) | 0.090 |
| *Myalgia/fatigue* | 433 (40.4%) | 420 (39.9%) | 13 (65.0%) | 0.042 |
| *Joint pain* | 21 (1.96%) | 20 (1.90%) | 1 (5.00%) | 0.329 |
| *Median pulse\*~* | 80.0 (76.0-87.0) | 80.0 (76.0-86.0) | 103 (80.0-116) | 0.020 |
| *>125 beats/min* | 8 (0.75%) | 7 (0.67%) | 1 (5.00%) | 0.086 |
| *Systolic pressure \** | 110 (110-120) | 112 (110-120) | 105 (100-130) | 0.252 |
| *<90 mmHg* | 11 (1.03%) | 8 (0.76%) | 3 (15.0%) | 0.001 |
| *Diastolic pressure \** | 76.0 (70.0-80.0) | 78.0 (70.0-80.0) | 65.0 (60.0-77.5) | 0.024 |
| *Respiratory rate \** | 18.0 (18.0-20.0) | 18.0 (18.0-20.0) | 23.0 (22.0-30.0) | <0.001 |
| *>24 breaths/min* | 25 (2.33%) | 20 (1.90%) | 5 (25.0%) | <0.001 |
| *SpO2* * | 97.0 (96.0-98.0) | 98.0 (96.0-98.0) | 86.0 (74.0-95.0) | <0.001 |
| **Laboratory findings** |  |  |  |  |
| White blood cells, $\times 10^9/L$, median | 6.30 (5.10-7.90) | 6.30 (5.07-7.90) | 7.05 (5.69-9.30) | 0.086 |
| $< 4 \times 10^9/L$ | 90 (8.8%) | 90 (9.0%) | 0 (0%) | 0.036 |
| $> 10 \times 10^9/L$ | 79 (7.8%) | 75 (7.5%) | 4 (22.2%) | |
| Neutrophil count, $\times 10^9/L$, median * | 3.80 (2.67-5.49) | 3.76 (2.66-5.47) | 4.96 (3.40-6.49) | 0.226 |
| $< 2 \times 10^9/L$ | 64 (13.3%) | 63 (13.4%) | 1 (8.3%) | 0.611 |
| Lymphocyte count, $\times 10^9/L$, median * | 1.73 (1.09-2.54) | 1.73 (1.10-2.54) | 1.76 (1.03-2.78) | 0.725 |
| $< 1 \times 10^9/L$ | 160 (23.3%) | 157 (23.3%) | 3 (25.0%) | 0.89 |
| NLR * | 2.06 (1.36-3.16) | 2.06 (1.37-3.14) | 2.37 (1.15-6.11) | 0.751 |
| $> 4.0$ | 76 (16.3%) | 72 (15.8%) | 4 (36.4%) | 0.068 |
| Median haemoglobin, g/L | 136 (124-150) | 137 (124-150) | 131 (119-142) | 0.200 |
| Monocytes, $\times 10^9/L$, median * | 0.38 (0.17-0.70) | 0.38 (0.18-0.70) | 0.23 (0.20-0.32) | 0.080 |
| Eosinophils, $\times 10^9/L$, median * | 2.00 (1.00-3.00) | 2.00 (1.00-3.00) | 2.00 (0.65-6.00) | 0.555 |
| Platelet count, $\times 10^9/L$, median * | 236 (196-288) | 236 (196-289) | 193 (154-241) | 0.008 |
| $< 100/L$ | 8 (0.8%) | 7 (0.7%) | 1 (5.9%) | 0.021 |
| Median prothrombin time, s * | 13.0 (11.7-16.0) | 13.0 (11.7-16.0) | 15.7 (12.6-18.5) | 0.063 |
| Median fibrinogen, g/L * | 3.10 (2.40-4.50) | 3.10 (2.40-4.32) | 5.90 (3.93-6.92) | 0.007 |
| Median albumin, g/L* | 40.0 (35.4-44.0) | 40.0 (35.5-44.0) | 30.5 (19.5-37.0) | 0.042 |
| Median alanine | 18.0 (9.78- | 18.0 (9.72-33.0) | 20.5 (12.1-42.5) | 0.459 |
| aminotransferase, U/L * | 33.0) |  |  |  |
| Median aspartate aminotransferase, U/L * | 18.5 (12.5-26.5) | 18.3 (12.4-25.9) | 42.0 (26.0-70.0) | <0.001 |
| Total bilirubin, mmol/L, median * | 11.2 (8.00-16.0) | 11.1 (8.00-16.0) | 12.9 (8.00-16.0) | 0.459 |
| Direct bilirubin, mmol/L, median * | 2.70 (1.79-4.30) | 2.62 (1.75-4.30) | 10.0 (4.00-10.9) | 0.012 |
| Median glucose, mmol/L * | 5.10 (4.50-6.00) | 5.10 (4.50-6.00) | 6.39 (5.16-9.30) | 0.002 |
| Median blood urea nitrogen, mmol/L * | 4.40 (3.47-5.54) | 4.40 (3.44-5.50) | 7.40 (4.74-15.4) | <0.001 |
| Median creatinine, uM* | 73.1 (60.3-88.0) | 73.0 (60.0-88.0) | 87.0 (75.8-163) | 0.002 |
| Median C-reactive protein, mg/L * | 2.30 (0.09-8.10) | 2.00 (0.02-7.90) | 8.30 (4.14-42.0) | 0.077 |
| Median sodium, mmol/L* | 141 (138-143) | 141 (138-143) | 136 (134-141) | 0.023 |
| Median potassium, mmol/L * | 4.10 (3.69-4.40) | 4.10 (3.69-4.40) | 3.80 (3.65-4.50) | 0.769 |
| **Radiologic findings \*** |  |  |  | 0.001 |
| Pneumonia | 230 (31.2%) | 218 (30.2%) | 12 (80.0%) |  |
| Bronchitis | 188 (25.5%) | 187 (25.9%) | 1 (6.67%) |  |
| **Treatments** |  |  |  |  |
| Antiviral medications | 749 (69.9%) | 739 (70.2%) | 10 (50.0%) | 0.087 |
| *Ribavirin* | 75 (7.00%) | 71 (6.75%) | 4 (20.0%) | 0.045 |
| *Lopinavir and ritonavir* | 696 (64.9%) | 688 (65.4%) | 8 (40.0%) | 0.034 |
| *Oseltamivir* | 30 (2.80%) | 29 (2.76%) | 1 (5.00%) | 0.436 |
| *Other* | 18 (1.7%) | 18 (1.7%) | 0 (0%) | 1 |
| Antibiotics | 308 (28.7%) | 299 (28.4%) | 9 (45.0%) | 0.170 |
| Hydroxychloroquine | 23 (2.15%) | 23 (2.19%) | 0 (0.00%) | 1.000 |
| Anticoagulant/antiplatelet | 29 (2.71%) | 24 (2.28%) | 5 (25.0%) | <0.001 |

| therapy |  |  |  |  |
| --- | --- | --- | --- | --- |
| Corticosteroids | 16 (1.49%) | 15 (1.43%) | 1 (5.00%) | 0.262 |
| Oxygen therapy | 64 (6%) | 44 (4.2%) | 20 (100%) | <0.001 |
| Mechanical ventilation | 27 (2.5%) | 9 (0.9%) | 18 (90%) | <0.001 |

Data are median (IQR) or n/N (%), where N is the number of patients with available data.
BMI=body mass index. COPD= Chronic obstructive pulmonary disease.
NLR=neutrophil/lymphocyte ratio. SpO2=oxygen saturation. \*Data were available for <1072 patients, see appendix p. 6 for details on available sample size.

Of all 1072 patients, 63·8 % had mild, 31·8% moderate, 3·83% severe 0·56% critical severity of disease on admission. A total of 20 deaths were reported: 1 (0·1%) in a patient with mild symptoms, 4 (1·2%) in patients with moderate disease, and 15 (31%) among patients with severe-critical disease (Table 1). The median age of the cohort was 36 years and 8·8 and 7·6% of the patients were younger than 15 and older than 65 years, respectively. Patients with moderate and severe-critical disease were older than those with mild disease by a median of 6 and 26 years, respectively (Table 1). Compared to survivors, non-survivors were older by a median of 30 years, with most exceeding 50 years of age, and were more likely to be male (Table 2).

Most patients (80%) were Kazakh; patients with severe-critical disease and non-survivors were less likely to be Kazakh, compared to non-severe patients (Table 1) and survivors (Table 2), respectively. The proportion of patients with comorbidities increased with disease severity, and 80% (16/20) of the non-survivors had a comorbidity. Presentation of most clinical symptoms and signs differed across disease severity categories (Table 1).

Compared to survivors, non-survivors were more likely to have fever, respiratory abnormalities, low blood pressure, and myalgia/fatigue on admission (Table 2). Disease severity was associated with altered markers of coagulation (prothrombin time, fibrinogen), liver (albumin, alanine aminotransferase (ALT), aspartate aminotransferase (AST), bilirubin) and kidney (blood urea nitrogen (BUN), creatinine) function, and changes in blood glucose, C-reactive protein and minerals (potassium, calcium). Severe-critical patients were also more likely to have elevated neutrophil-to-lymphocyte ratios (33% of the patients) compared to mild and moderate severity patients (16 and 14%, respectively). Compared to survivors, non-survivors were more likely to have leukocytosis (7·5 vs 22% of the patients, respectively), lower platelet counts and altered fibrinogen, albumin, AST, bilirubin, glucose, BUN, creatinine, and sodium levels. D-dimer and lactate measurements were excluded from the analysis due to the small proportion of patients with available data. Chest X-ray and/or computed tomography (CT) data were available for 418 (39%) patients, of whom 57% had abnormalities defined as “pneumonia”(31%) or “bronchitis”(26%) by the radiologists in charge. Pneumonia was most frequently diagnosed in severe-critical patients (91%, Table 1) and non-survivors (80%, Table 2) compared to patients with non-severe disease and survivors.

Antibiotics and anticoagulant or antiplatelet therapy were more likely administered to patients with severe-critical disease, while significantly fewer patients with severe-critical disease received antiviral medications compared to patients with non-severe disease (Table 1). Compared with survivors, non-survivors were more likely to receive ribavirin and lopinavir-ritonavir and anticoagulant/antiplatelet therapy (Table 2). Oxygen therapy was administered to a total of 64 (6%) patients, chiefly those with severe-critical disease (65%) and all non-survivors (100%). Mechanical ventilation was performed on 27 (3%) patients, of whom 40% had severe-critical disease and 90% were non-survivors (Tables 1 and 2).

In the univariable analysis of COVID-19 severity predictors, older age, non-Kazakh ethnicity, comorbidities, elevated white blood cells (WBC), high neutrophil-to-lymphocyte ratio (NLR), lower haemoglobin, lower albumin and elevated creatinine were all associated with severe disease. We included 1072 patients (1024 non-severe and 47 severe-to-critical patients) in the multivariable analysis, with two continuous and two categorical variables (age, ethnicity, comorbidities, and white blood cells (WBC). Older age (OR 1·05 [95% CI 1·03-1·07], p<0.001), comorbidities (OR 2·13 [95% CI 1·07-4·23], p<0·031) and elevated WBC (OR 1·14 [95% CI 1·01-1·28], p<0·032) were associated with increased odds of severe disease (Table 3).

**Table 3:** Bivariate logistic regression of factors associated with COVID-19 disease severity on admission.

|  | **Univariable OR**<br>**(95% CI)** | **p value** | **Multivariable**<br>**OR (95% CI)** | **p value** |
| --- | --- | --- | --- | --- |
| Age, years* | 1.06 (1.04-1.08) | <0.001 | 1.05 (1.03-1.07) | <0.001 |
| Male sex (vs. female) | 1.74 (0.97-3.14) | 0.063 | .. | .. |
| Kazakh ethnicity (vs. other ethnicities) | 0.41 (0.22-0.75) | 0.004 | 0.78 (0.39-1.57) | 0.484 |
| Comorbidities | 3.46 (1.86-6.45) | <0.001 | 2.13 (1.07-4.23) | 0.031 |
| White blood cells* | 1.13 (1.01-1.26) | 0.035 | 1.14 (1.01-1.28) | 0.032 |
| NLR> 4.0 | 2.78 (1.2-6.46) | 0.017 | .. | .. |
| Haemoglobin, g/L* | 0.98 (0.97-0.99) | 0.002 | .. | .. |
| Prothrombin time, s* | 1 (0.999-1) | 0.746 | .. | .. |
| Fibrinogen, g/L* | 0.998 (0.99-1.01) | 0.689 | .. | .. |
| Albumin, g/L* | 0.94 (0.91-0.98) | 0.001 | .. | .. |
| Aspartate aminotransferase, U/L* | 1 (1-1) | 0.761 | .. | .. |
| Total bilirubin, mmol/L* | 1 (0.999-1) | 0.804 | .. | .. |
| Glucose, mmol/L* | 1 (0.999-1.0) | 0.807 | .. | .. |
| Blood urea nitrogen, mmol/L* | 1 (0.999-1.0) | 0.846 | .. | .. |
| Creatinine, uM* | 1.01 (1.0-1.01) | 0.004 | .. | .. |
| C-reactive protein, mg/L* | 1 (1-1) | 0.843 | .. | .. |
| Potassium, mmol/L* | 0.87 (0.58-1.30) | 0.486 | .. | .. |
| Calcium, mmol/L* | 1.04 (0.98-1.11) | 0.208 | .. | .. |

The two binary dependent variables in the model were patients with “severe”(47 patients) versus “non-severe” (1024 patients) COVID-19 disease. OR=odds ratio. NLR=neutrophil/lymphocyte ratio. \*For each additional unit.

In the univariable analysis of COVID-19 mortality predictors, older age, male sex, non-Kazakh ethnicity, comorbidities, elevated WBC, decreased platelet counts, lower albumin and elevated creatinine were associated with death. 1072 patients (1052 survivors and 20 non-survivors) were included in the multivariable analysis and a backward elimination strategy was used to identify variables describing the best model. Older age (OR 1·09 [95% CI 1·06-1·12], p<0·001) and male sex (OR 5·97 [95% CI 1·95-18·32], p<0·002) were associated with increased odds of death (Table 4).

**Table 4:** Bivariate logistic regression of factors associated with COVID-19 mortality in Kazakhstan.

|  | **Univariable OR**<br>**(95% CI)** | **p**<br>**value** | **Multivariable**<br>**OR (95% CI)** | **p value** |
| --- | --- | --- | --- | --- |
| Age, years* | 1.08 (1.05-1.11) | <0.001 | 1.09 (1.06-1.12) | <0.001 |
| Sex (Men) | 2.89 (1.1-7.58) | 0.031 | 5.97 (1.95-18.32) | 0.002 |
| Kazakh ethnicity | 0.25 (0.10-0.61) | 0.002 | .. | .. |
| Comorbidities | 6.14 (2.04-18.49) | 0.001 | .. | .. |
| White blood cells, $\times 10^9/L^*$ | 1.22 (1.04-1.43) | 0.014 | .. | .. |
| Platelet count, $\times 10^9/L^*$ | 0.99 (0.98-0.995) | 0.002 | .. | .. |
| Fibrinogen, g/L* | 0.999 (0.99-1.01) | 0.745 | .. | .. |
| Albumin, g/L* | 0.93 (0.88-0.98) | 0.004 | .. | .. |
| Aspartate aminotransferase, U/L* | 1 (1-1) | 0.839 | .. | .. |
| Direct bilirubin, mmol/L* | 1 (0.998-1.00) | 0.916 | .. | .. |
| Glucose, mmol/L* | 1.04 (0.989-1.09) | 0.132 | .. | .. |
| Blood urea nitrogen, mmol/L* | 1 (0.999-1.00) | 0.865 | .. | .. |
| Creatinine, $\mu M^*$ | 1.01 (1.00-1.01) | 0.001 | .. | .. |
| Sodium, mmol/L* | 0.998 (0.97-1.03) | 0.884 | .. | .. |

The two binary dependent variables in the model were “non-survivors” (20 patients) versus “survivors” (1052 patients). OR=odds ratio. NLR=neutrophil/lymphocyte ratio. \*For each additional unit.

Genomic data were generated for the viral samples acquired from all 53 patients (median age 21 years (IQR 21-34), 66% male, 94% Kazakh, see Fig 1B and appendix p. 5). Compared to the Wuhan-1 reference, there were a maximum of 12 single-nucleotide polymorphisms (SNPs) per virus genome (median 9 SNPs, IQR 8-10, see appendix p. 6), corroborating little variation observed among the SARS-CoV-2 strains ^15,16^. The Kazakhstan (Kaz) strains grouped into 7 distinct global lineages: O/B.4.1 (n=27), S/A.2 and S/B.1.1 (19), GH/B.1.255 and GH/B.1.3 (n=5), G/B.1 (n=1) and GR/B.1.1.10 (n=1) (Fig 4A, appendix p.6). Kaz_O/B4.1 isolates clustered with predominantly Middle Eastern sequences that also included European isolates (Fig 4B), Kaz_S/A2-B.1.1 viruses were closely related to the strains from Spain, Britain, and Russia, although this clade appears to have predominantly Asian origins (Fig 4B). Kaz_GH/B.1.255-B.1.3 viruses were nested with Mexican and Argentinian viruses deriving ancestral lineages from both the Middle East and Europe. Kaz_G/B.1 and Kaz_GR/B.1.1.10 viruses cluster closely with North-Western/Western European and Southern European viruses, respectively (Fig 4B).

**Figure 4:**
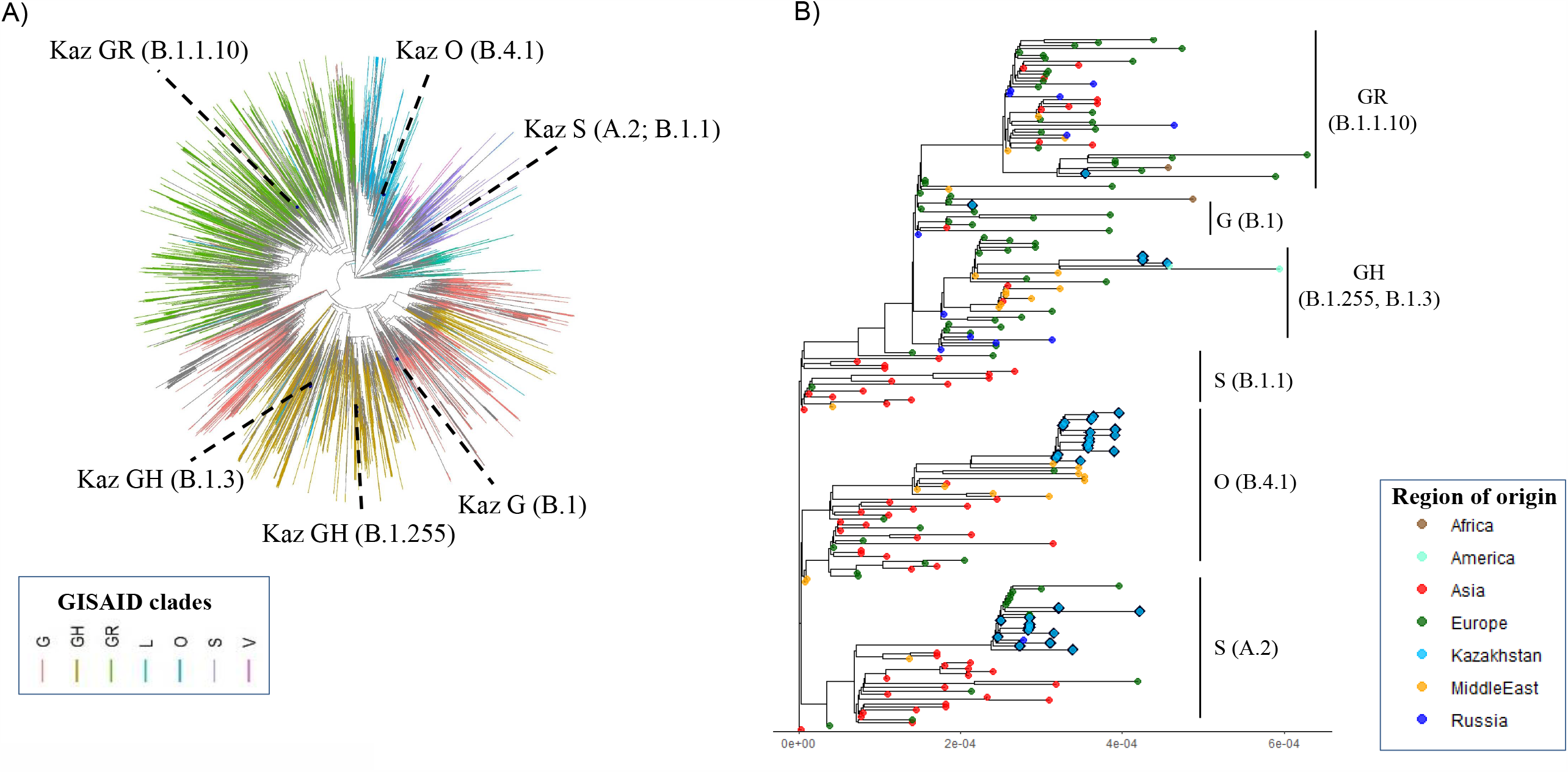
**A**. Phylogenetic tree depicting the Kazakhstan (“Kaz”) virus isolates in the context of globally circulating SARS-CoV-2 lineages. Each clade is denoted by a corresponding Global Initiative on Sharing All Influenza Data (GISAID) clade name; the Pangolin lineage names are given in brackets. Branch lengths measured in units of substitutions per site. Tree is coloured based on the GISAID nomenclature (see legend). **B**. Phylodynamic analysis of the Kazakhstan SARS-CoV-2 sequences in the international context. Maximum likelihood tree of Kazakhstan viral sequences and a subset of international sequences (see Methods), coloured by region of origin.

## DISCUSSION

Here we detailed the clinical features of 1072 patients with laboratory confirmed COVID-19 and described the genetic diversity of circulating SARS-CoV-2 in Kazakhstan. Consistent with data from other cohorts^1,17–22^, older age, comorbidities, and elevated WBC on admission were associated with higher odds of severe disease, while higher odds of death were associated with older age and male sex. Additional factors observed in patients with severe disease included non-Kazakh ethnicity, high NLR, lower haemoglobin, lower albumin and elevated creatinine. Other factors commonly observed in non-survivors included non-Kazakh ethnicity, comorbidities, elevated WBC, decreased platelet counts, lower albumin and elevated creatinine. Our genomic findings point at several independent importations of SARS-CoV-2, followed by community-level amplification early in the pandemic.

A highlight of the current cohort is a lower ratio of patients with severe-critical to non-severe (moderate and mild) COVID-19 compared to other cohorts, despite an early implementation in Kazakhstan of the WHO disease classification guidelines^13^. Thus, the proportion of severe-critical cases in our study was 4%, which is approximately 5-fold lower than that reported elsewhere ^17,21,23^. At the same time, the overall mortality rate in our cohort (1.9%) is consistent with the estimates of crude confirmed case fatality risk among COVID-19 patients presenting a wide spectrum of disease manifestations ^24^, but is 10-fold lower than the rates typically described for hospitalized patients ^25,26^-probably because in Kazakhstan all laboratory-confirmed COVID-19 patients, regardless of disease manifestation on admission, were admitted to hospitals. As expected, in our cohort mortality correlated with disease severity; the risk of death was low in patients with mild disease, similar to that reported typically (∼0.1%) ^17^, but it escalated dramatically to a 4-fold higher risk in patients with severe-critical disease compared to severe COVID-19 elsewhere (e.g. 8% in ^17^). The high mortality associated with severe-critical COVID-19 in this cohort could be due to a combination of mis-categorization of patients manifesting severe COVID-19 symptoms into non-severe disease categories, and subsequent “saturation” of the severe-critical disease category by patients with unfavorable prognosis, and a shortage of healthcare resources in a rapidly escalating pandemic ^2,3^.

Consistent with other studies, including those from the neighbouring Uzbekistan and Russia ^1,17–20,24,27^, older age was a key contributor to COVID-19 severity and mortality, and the non-survivor group consisted predominantly of older men. The cohort’s median age (36 years) was higher than the Kazakhstan median (31 years)^28^ and identical to that of COVID-19 patients in Uzbekistan ^27^. The cohort sex ratio and ethnic make-up were consistent with the recent census data ^28,29^, whereby most Kazakhstan residents are Kazakh (68·5%), followed by peoples of Russian (18·9%) and of other ancestries (12·6%), principally Uzbeks, Uyghurs, Ukrainians, Tatars and Germans. In univariable analyses, non-Kazakh ethnicity was associated with both COVID-19 severity and death, although this effect was abrogated in multivariable models, suggesting that non-Kazakh ethnicity is linked to other risk factors, consistent with augmented risk of COVID-19 infection and more severe disease among ethnic minority groups in Western countries ^30^. A biological predisposition of non-Kazakh people to a more severe COVID-19 disease should be addressed by future investigations.

Overall, both our clinical and laboratory findings were in line with earlier research. Notwithstanding, there are differences that may have arisen due to the heterogeneity in data collection by facilities across the country. For example, unlike in other studies ^17,22^, thrombocytopenia in our cohort was uncommon, but, consistently, lower absolute platelet counts were associated with death. Further, differences in other blood count parameters were not significant among the patient sub-categories, but consistent changes associated with COVID-19 severity were observed in blood biochemistry, coagulation and liver and renal function tests, inflammatory markers and electrolytes ^17,22,25^. These observations emphasize the need for careful re-examination of the diagnostic test reference ranges utilized across the country in the context of COVID-19 patient management. Predictably, the prevalence of radiology-confirmed pneumonia increased with disease severity; based on radiological assessment, bronchitis was a common diagnosis in non-severe COVID-19, consistent with bronchial wall thickening in ∼20% of COVID-19 cases, indicating an effect of SARS-CoV-2 with or without a respiratory coinfection^31^.

We identified representatives of five of the eight global SARS-CoV-2 clades circulating in Kazakhstan early in the pandemic. Most isolates (46/53) belonged to clades O or S - descendants of the early lineages that originated in Asia and were globally prevalent prior to the appearance of the D614-G614 mutation that swept through Europe, reaching a ∼67% frequency among European sequences by mid-March ^32^. Since our viral isolates were sampled after the appearance of the G clade mutation in Europe, the high prevalence of O and S sub-types in Kazakhstan suggests an early importation, perhaps weeks prior to the declaration of the international travel ban, and subsequent amplification through community spread. Notably, the Kazakhstan S lineage isolates clustered with viruses of from Europe, such as Spain in the case of lineage S/A.2, while the Kazakhstan O clade isolates, clustering with Middle Eastern (particularly, Iranian) strains, all belonged to lineage B.4.1, which arose uniquely in Kazakhstan. These observations are consistent with the evidence that areas of southern Europe and the Middle East represented early points of SARS-CoV-2 introduction and spread ^33^. Additionally, the presence of three clade G lineages in Kazakhstan at diverse timepoints, between 25 March and 9 May, indicates multiple independent importations from Europe and the Americas.

Patient travel histories suggest that during the study period COVID-19 infections were imported mainly from the neighboring Russia, and from Europe. Only 1% of the patients reported travel from China, where case numbers had substantially decreased by March 2020 ^34^. Remarkably, 41% of the international travelers used train or bus, and other means of ground transportation, which has implications for public health policies aimed to curtail COVID-19 spread on public transit.

Our study has several limitations. First, our cohort represents only a third of all COVID-19 cases confirmed by PCR in March-April in Kazakhstan. Second, the quality of our dataset is largely dependent on the quality of the medical record data, which were collected in emergency settings by clinicians and which could have increased data heterogeneity. Third, some data, such as laboratory findings for CRP, were only available for a subset of participants, while other data, such as D-dimer levels and information on clinical outcomes or complications were available for <40% of patients within the comparison groups and were excluded from the analyses. Fourth, death reporting may have been delayed or more deaths may have occurred after April 30, resulting in under-estimation of mortality. Fifth, laboratory confirmation of COVID-19 was done using a commercially available PCR supplied centrally to all testing sites, but we were unable to access data on the intra- and inter-laboratory performance of this test. Sixth, our genomic analysis was limited to a qualitative assessment and cannot fully recapitulate the country-wide SARS-CoV-2 diversity due to the small sample size and geographic constraints.

To conclude, we are hopeful that these clinical and virologic data on COVID-19 will help shape public health policies and therapeutic and vaccine interventions in Kazakhstan. Our findings could ultimately be applicable to other neighbouring countries that are similar in their ethnic and socio-cultural profiles and healthcare structures, meriting consideration by a broad spectrum of international public health authorities and policy makers.

## Data Availability

The retrospective cohort study was approved by the Research Ethics Board of Semey Medical University as an anonymized epidemiological study, for which the requirement for informed consent was waived due to the pandemic state and urgent need to collect and analyze data. Virological samples used in genomic studies were anonymized and all genomic study procedures were approved by the Ethics Committee of the National Centre for Biotechnology (Nur-Sultan, Kazakhstan).

https://www.gisaid.org/

## DECLARATIONS

### Contributors

SY, MG and RI: conceived, designed, and implemented the study, drafted the manuscript. SY, MG, RI: analyzed the patient data. SY: performed statistical analyses and contributed to genomic analyses. SVG, DB: contributed to statistical analyses and performed genomic analyses. AS and the CGRG group performed sequencing experiments and did initial genomic data analysis. KSM provided laboratory advice, and clinical guidance and contributed to data analysis. the SCERG group extracted data from electronic medical records. RI, YZ: facilitated data acquisition, and supervised the study. All authors contributed to data interpretation, critically reviewed the manuscript draft, and approved the final version for submission.

## Acknowledgements

We thank all the participants, public health, and clinical and laboratory staff, who have been involved in COVID-19 diagnostic testing, clinical care and case management in Kazakhstan. The authors acknowledge the contributions from other laboratories to GISAID (see appendix pp. 10-22).

## Data sharing

The datasets from this study will be made available, wherever possible, when not otherwise restricted by ongoing collaborative research, on appropriate request to the corresponding author. SARS-CoV-2 genomic data have been submitted to GISAID under accession IDs# EPI_ISL_435045-435048; EPI_ISL_454497-454520; EPI_ISL_454571-454604.

## Supplementary information

All supplementary information can be found in the Appendix.

## References

1 Hu B, Guo H, Zhou P, Shi Z-L. Characteristics of SARS-CoV-2 and COVID-19. Nat Rev Microbiol 2020; published online Oct 6. DOI:10.1038/s41579-020-00459-7.

2 Hopman J, Allegranzi B, Mehtar S. Managing COVID-19 in Low- and Middle-Income Countries. JAMA 2020; 323: 1549–50.

3 Balakrishnan VS. COVID-19 response in central Asia. The Lancet Microbe 2020; 1: e281.

4 Semenova Y, Glushkova N, Pivina L, et al. Epidemiological Characteristics and Forecast of COVID-19 Outbreak in the Republic of Kazakhstan. J Korean Med Sci 2020; 35. DOI:10.3346/jkms.2020.35.e227.

5 Bayesheva D, Boranbayeva R, Turdalina B, et al. COVID-19 in the paediatric population of Kazakhstan. Paediatr Int Child Health 2020; : 1–7.

6 Hadfield J, Megill C, Bell SM, et al. Nextstrain: real-time tracking of pathogen evolution. Bioinformatics 2018; 34: 4121–3.

7 Bogner P, Capua I, Lipman DJ, Cox NJ. A global initiative on sharing avian flu data. Nature 2006; 442: 981–981.

8 Seemann T. Source code for snp-dists software. 2018; published online Sept 9. DOI:10.5281/zenodo.1411986.

9 Katoh K, Standley DM. MAFFT multiple sequence alignment software version 7: improvements in performance and usability. Mol Biol Evol 2013; 30: 772–80.

10 Nguyen L-T, Schmidt HA, von Haeseler A, Minh BQ. IQ-TREE: a fast and effective stochastic algorithm for estimating maximum-likelihood phylogenies. Mol Biol Evol 2015; 32: 268–74.

11 Yu G. Using ggtree to Visualize Data on Tree-Like Structures. Curr Protoc Bioinformatics 2020; 69: e96.

12 Rambaut A, Holmes EC, O’Toole Á, et al. A dynamic nomenclature proposal for SARS-CoV-2 lineages to assist genomic epidemiology. Nat Microbiol 2020; 5: 1403–7.

13 Clinical management of COVID-19. https://www.who.int/publications-detail-redirect/clinical-management-of-covid-19 (accessed Dec 14, 2020).

14 Kazakhstan: WHO Coronavirus Disease (COVID-19) Dashboard. https://covid19.who.int (accessed Dec 14, 2020).

15 Seemann T, Lane CR, Sherry NL, et al. Tracking the COVID-19 pandemic in Australia using genomics. Nat Commun 2020; 11: 4376.

16 Lu R, Zhao X, Li J, et al. Genomic characterisation and epidemiology of 2019 novel coronavirus: implications for virus origins and receptor binding. Lancet 2020; 395: 565–74.

17 Guan W-J, Ni Z-Y, Hu Y, et al. Clinical Characteristics of Coronavirus Disease 2019 in China. N Engl J Med 2020; 382: 1708–20.

18 Munblit D, Nekliudov NA, Bugaeva P, et al. StopCOVID cohort: An observational study of 3,480 patients admitted to the Sechenov University hospital network in Moscow city for suspected COVID-19 infection. Clin Infect Dis 2020; published online Oct 9. DOI:10.1093/cid/ciaa1535.

19 Demkina AE, Morozov SP, Vladzymyrskyy AV, et al. Risk factors for outcomes of COVID-19 patients: an observational study of 795 572 patients in Russia. medRxiv 2020; : 2020.11.02.20224253.

20 Moiseev S, Avdeev S, Brovko M, Bulanov N, Tao E, Fomin V. Outcomes of intensive care unit patients with COVID-19: a nationwide analysis in Russia. Anaesthesia 2020; published online Oct 5. DOI:10.1111/anae.15265.

21 Wu Z, McGoogan JM. Characteristics of and Important Lessons From the Coronavirus Disease 2019 (COVID-19) Outbreak in China: Summary of a Report of 72L314 Cases From the Chinese Center for Disease Control and Prevention. JAMA 2020; 323: 1239–42.

22 Zhou F, Yu T, Du R, et al. Clinical course and risk factors for mortality of adult inpatients with COVID-19 in Wuhan, China: a retrospective cohort study. Lancet 2020; 395: 1054–62.

23 Tabata S, Imai K, Kawano S, et al. Clinical characteristics of COVID-19 in 104 people with SARS-CoV-2 infection on the Diamond Princess cruise ship: a retrospective analysis. Lancet Infect Dis 2020; 20: 1043–50.

24 Wu JT, Leung K, Bushman M, et al. Estimating clinical severity of COVID-19 from the transmission dynamics in Wuhan, China. Nat Med 2020; 26: 506–10.

25 Richardson S, Hirsch JS, Narasimhan M, et al. Presenting Characteristics, Comorbidities, and Outcomes Among 5700 Patients Hospitalized With COVID-19 in the New York City Area. JAMA 2020; 323: 2052–9.

26 RECOVERY Collaborative Group, Horby P, Lim WS, et al. Dexamethasone in Hospitalized Patients with Covid-19 - Preliminary Report. N Engl J Med 2020; published online July 17. DOI:10.1056/NEJMoa2021436.

27 Kim K, Choi JW, Moon J, et al. Clinical Features of COVID-19 in Uzbekistan. J Korean Med Sci 2020; 35. DOI:10.3346/jkms.2020.35.e404.

28 | Human Development Reports. http://hdr.undp.org/en/countries/profiles/KAZ (accessed Dec 14, 2020).

29 Agency for Strategic planning and Reforms of the Republic of Kazakhstan, Bureau of National Statistics. https://www.stat.gov.kz/ (accessed Dec 14, 2020).

30 Sze S, Pan D, Nevill CR, et al. Ethnicity and clinical outcomes in COVID-19: A systematic review and meta-analysis. EClinicalMedicine 2020; : 100630.

31 Carotti M, Salaffi F, Sarzi-Puttini P, et al. Chest CT features of coronavirus disease 2019 (COVID-19) pneumonia: key points for radiologists. Radiol Med 2020; : 1–11.

32 Korber B, Fischer WM, Gnanakaran S, et al. Tracking Changes in SARS-CoV-2 Spike: Evidence that D614G Increases Infectivity of the COVID-19 Virus. Cell 2020; 182: 812- 827.e19.

33 Lemey P, Hong SL, Hill V, et al. Accommodating individual travel history and unsampled diversity in Bayesian phylogeographic inference of SARS-CoV-2. Nat Commun 2020; 11: 5110.

34 Leung K, Wu JT, Liu D, Leung GM. First-wave COVID-19 transmissibility and severity in China outside Hubei after control measures, and second-wave scenario planning: a modelling impact assessment. Lancet 2020; 395: 1382–93.

