## Appendix for "Epidemiological and Clinical Characteristics, and Virologic Features of COVID-19 Patients in Kazakhstan: a Nation-Wide, Retrospective, Cohort Study"

### SUPPLEMENTARY MATERIAL

**Whole genome sequencing methods**

**Sample collection:** Nasopharyngeal swabs were collected from 53 symptomatic patients during clinical assessment. Samples were transported at 4-8C in transport medium (medium 199 with Hanks' salts (Sigma, M0393), 4 ml Gentamicin solution (Sigma, G 1397), 11 ml BSA, fraction V (Sigma, A-8412), 11 ml 1 M HEPES buffer solution (Sigma, 83264) and delivered to the laboratory within 8 hours of collection.
**SARS-CoV-2 infection validation:** A commercially available Real-Time Fluorescent RT-PCR kit (BGI, Shenzhen) was used to confirm SARS-CoV-2-positive samples. For the NGS sequencing, samples were selected up to the 24th PCR threshold cycle.
**RNA extraction and amplification:** Total RNA was isolated using the GeneJet Viral DNA and RNA Purification Kit (Thermo Scientific) and quantified by fluorometry. Reverse transcription was performed with three primer mixes (see Table in appendix p. 3) and SuperScript™ IV Reverse Transcriptase (Invitrogen™) in a 40 µL reaction mixture containing 1x SSIV Buffer, 0,5 mM each dNTP, 5 mM DTT, 100 nM each primer, 10 U/µL SuperScript® IV Reverse Transcriptase and 20 µL RNA. Incubated at 50 °C for 30 minutes. The viral genome was amplified in 30 fragments, 944 -1364 bp long and an overlap of at least 170 bp. (see Table, appendix p. 4). Primers were designed using the FastPCR software [1] using the Wuhan-1 sequence (Genbank accession number NC_045512) as reference. Each fragment was amplified in a singleplex PCR reaction with a volume of 25 µl. The reaction mixture included: 1× Platinum™ II PCR Buffer, 0,2 mM each dNTP dNTP, 200 nM forward and reverse primer, 0.04 U/µL Platinum™ II Taq Hot-Start DNA Polymerase, 3 µL cDNA. PCR amplification program included: long-term denaturation at 94 ° C for 2 minutes; 10 cycles: 94 °C - 15 seconds, 58 °C - 30 seconds, 68 °C - 45 seconds; 25 cycles: 94 °C - 15 seconds, 58 °C - 15 seconds, 68 °C - 45 seconds, final elongation 15 minutes at 68 °C.
**Sequencing strategy:** Four samples (NCB1-5, see appendix p. 5) were sequenced by the Sanger dideoxy sequencing approach. PCR products were purified with ExoSAP-IT ™ (Applied Biosystems) and sequenced using the BigDye® Terminator v3.1 Cycle Sequencing Kit (Applied Biosystems) and primers used for PCR amplification. Separation was performed using a 3730xl DNA Analyzer (Applied Biosystems). Collection of sequences was carried out using the SeqScape Software v. 3.0 according to the Wuhan-1 reference.
The rest 49 samples were sequenced using next generation Illumina sequencing technology as follows. PCR products were mixed into one mixture, which was purified using 0.6 volume AMPure XP (Beckman Coulter) and used to prepare libraries using Nextera DNA Flex Library Prep Kit (Illumina, USA), according to the manufacturer's instructions. Sequencing was performed on a MiSeq sequencer (Illumina, USA) using the MiSeq Reagent Kit v3, 600 Cycles (Catalog # MS-102-3003).
**Virus genome analysis and annotation:** Raw data quality control was carried out using FastQC v0.11.7 [2] MultiQC v1.8 [3]. Raw data trimming was carried out using the Seqkit v1.3-r106 software [4]. FASTQ sequences were aligned to the Wuhan-1 reference using using the BWA v0.7.17-r1188 software [5]. For multiple genome alignment, fasta text files were imported into Ugene (http://ugene.net/) and the Illumina BaseSpace DRAGEN Pathogen Detection software with the following parameters: somatic small variant base caller, k-mers generated from the Wuhan-1 reference, minimum depth=10, minimum allele frequency=0.5, virus detection threshold 5% of the genome at 5X coverage. Variants were identified and a consensus sequence was obtained using Samtools v1.10 [6] и Bcftools v1.10.2-54 [7].

### Primers used in RT-PCR

| | Primer name | Sequencing 5'-3' |
|---|---|---|
| **Primers#1** | | |
| | **Primer name** | **Sequencing 5'-3'** |
| | SARS-Cov2-RT-03 | CAGTTGTGATGATTCCTAAGAAAACAA |
| | SARS-Cov2-RT-09 | CTGAACTCACTTTCCATCCAAC |
| | SARS-Cov2-RT-12 | GAATTTGTCTGCTAATAATGCAGC |
| | SARS-Cov2-RT-15 | CGTTCAATCATAAGTGTACCATC |
| | SARS-Cov2-RT-18 | CCAGCTTGTAGACGTACTGT |
| | SARS-Cov2-RT-20N | GAAAGTAACACCTGAGCATTGTC |
| | SARS-Cov2-RT-23 | GTGTTATTAGCTCTCAGGTTGTC |
| | SARS-Cov2-RT-26 | TAGGTGGAATGTGGTAGGATTAC |
| | SARS-Cov2-RT-29 | CGCACAGAATTTTGAGCAGTTTC |
| | SARS-Cov2-RT-30 | CCCTCTTAGTGTCAATAAAGTCC |
| **Primers#2** | | |
| | SARS-Cov2-RT-01 | tttttttttttgtcattctcctaagaagcta |
| | SARS-Cov2-RT-04 | ACAGCTCCGATTACGAGTTC |
| | SARS-Cov2-RT-06 | AAGGATCATAAACTGTGTTGTTGAC |
| | SARS-Cov2-RT-10 | GTACAGTTGCACAATCACCAATC |
| | SARS-Cov2-RT-13 | TTGTAAAGTTGCCACATTCCTAC |
| | SARS-Cov2-RT-16 | GTGCATCTTGATCCTCATAACTC |
| | SARS-Cov2-RT-19 | TACAGCACCCTGCATGGAAA |
| | SARS-Cov2-RT-22 | CCACTTCTCTTGTTATGACTGC |
| | SARS-Cov2-RT-24 | AGTCGGCATAGATGCTTTAATTC |
| | SARS-Cov2-RT-28 | GCCATACTCCACTCATCTAAATC |
| **Primers#3** | | |
| | SARS-Cov2-RT-02 | CATTCTAGCAGGAGAAGTTCC |
| | SARS-Cov2-RT-05 | TTCCCATTTTTCAGTATAACCACC |
| | SARS-Cov2-RT-07 | CTCTTGCTTGGTTTTGATGGATC |
| | SARS-Cov2-RT-08 | CACCATTACAAGGTGTGCTAC |
| | SARS-Cov2-RT-11 | ACTTGACCATCAACTCTACCATC |
| | SARS-Cov2-RT-14 | CTAACATAGTGCTCTTGTGGC |
| | SARS-Cov2-RT-17 | GCATGGCATCACAGAATTGTAC |
| | SARS-Cov2-RT-21N | TTGCGTAAGAGGTAATAGCACATC |
| | SARS-Cov2-RT-25 | GTAAAGCACCGTCTATGCAATAC |
| | SARS-Cov2-RT-27 | TTTTCTCTGTTCAACTGAAGGTTTAC |

**Primers used for SARS-CoV-2 genome amplification and NGS sequencing**

| Forward primer | Sequencing 5'-3' | Reverse primer | Sequencing 5'-3' |
|---|---|---|---|
| SARS-Cov2-RT-01 | ttttttttttttgtcattctcctaagaagcta | SARS-Cov2-PCR-35 | CCCTATGGTGCTAACAAAGAC |
| SARS-Cov2-RT-02 | CATTCTAGCAGGAGAAGTTCC | SARS-Cov2-PCR-34 | CAGACAAGAGGAAGTTCAAGAAC |
| SARS-Cov2-RT-03 | CAGTTGTGATGATTCCTAAGAAAACAA | SARS-Cov2-PCR-33 | CAGTAACTTTAGCTTGTTTTGTGC |
| SARS-Cov2-RT-04 | ACAGCTCCGATTACGAGTTC | SARS-Cov2-PCR-32 | GCTTTAGTCTACTTCTTGCAGAG |
| SARS-Cov2-RT-05 | TTCCCATTTTTCAGTATAACCACC | SARS-Cov2-PCR-31N | CCCTCAGTCAGCACCTCA |
| SARS-Cov2-RT-06 | AAGGATCATAAACTGTGTTGTTGAC | SARS-Cov2-PCR-30 | CTATTAGTGTTACCACAGAAATTCTAC |
| SARS-Cov2-RT-07 | CTCTTGCTTGGTTTTGATGGATC | SARS-Cov2-PCR-29 | GAGGTGATGAAGTCAGACAAATC |
| SARS-Cov2-RT-08 | CACCATTACAAGGTGTGCTAC | SARS-Cov2-PCR-28 | TGGTTCCATGCTATACATGTCTC |
| SARS-Cov2-RT-09 | CTGAACTCACTTTCCATCCAAC | SARS-Cov2-PCR-23 | ATGCTATTAGAAAAGTGTGACCTTC |
| SARS-Cov2-RT-10 | GTACAGTTGCACAATCACCAATC | SARS-Cov2-PCR-21 | TTACCTGTTAATGTAGCATTTGAGC |
| SARS-Cov2-RT-11 | ACTTGACCATCAACTCTACCATC | SARS-Cov2-PCR-20 | CAGGTAACCTACAAAGCAACCA |
| SARS-Cov2-RT-12 | GAATTTGTCTGCTAATAATGCAGC | SARS-Cov2-PCR-19 | GAGAAAAGCTGTCTTTATTTCACC |
| SARS-Cov2-RT-13 | TTGTAAAGTTGCCACATTCCTAC | SARS-Cov2-PCR-18 | TGTCTTATGGTATTGCTACTGTAC |
| SARS-Cov2-RT-14 | CTAACATAGTGCTCTTGTGGC | SARS-Cov2-PCR-17 | TTGTGTGTTTCAATAGCACTTATGC |
| SARS-Cov2-RT-15 | CGTTCAATCATAAGTGTACCATC | SARS-Cov2-PCR-16N | CTATGACTTTGCTGTGTCTAAGG |
| SARS-Cov2-RT-16 | GTGCATCTTGATCCTCATAACTC | SARS-Cov2-PCR-15 | TAGAATAGACGGTGACATGGTAC |
| SARS-Cov2-RT-17 | GCATGGCATCACAGAATTGTAC | SARS-Cov2-PCR-14 | GTACTACACAAACTGCTTGCAC |
| SARS-Cov2-RT-18 | CCAGCTTGTAGACGTACTGT | SARS-Cov2-PCR-13 | AAGAATAGCATAGATGCCTTCAAAC |
| SARS-Cov2-RT-19 | TACAGCACCCTGCATGGAAA | SARS-Cov2-PCR-12 | TTTAACCTTGTGGCTATGAAGTAC |
| SARS-Cov2-RT-20N | GAAAGTAACACCTGAGCATTGTC | SARS-Cov2-PCR-11 | ATGGTTATGTTCACACCTTTAGTAC |
| SARS-Cov2-RT-21N | TTGCGTAAGAGGTAATAGCACATC | SARS-Cov2-PCR-10N | CACCTCTTCATGTCATGTCTAAAC |
| SARS-Cov2-RT-22 | CCACTTCTCTTGTTATGACTGC | SARS-Cov2-PCR-09N | GTGTTAATTGTGATACATTCTGTGCT |
| SARS-Cov2-RT-23 | GTGTTATTAGCTCTCAGGTTGTC | SARS-Cov2-PCR-08 | TGCTTATGTAGACAATTCTAGTCTTAC |
| SARS-Cov2-RT-24 | AGTCGGCATAGATGCTTTAATTC | SARS-Cov2-PCR-07 | AGGGTGTAGAAGCTGTTATGTAC |
| SARS-Cov2-RT-25 | GTAAAGCACCGTCTATGCAATAC | SARS-Cov2-PCR-06 | CAAAGTGCCAGCTACAGTTTC |
| SARS-Cov2-RT-26 | TAGGTGGAATGTGGTAGGATTAC | SARS-Cov2-PCR-05 | TCAGCACGAAGTTCTACTTGC |
| SARS-Cov2-RT-27 | TTTTCTCTGTTCAACTGAAGGTTTAC | SARS-Cov2-PCR-04 | ACAATACCTTCACACTCAAAGGC |
| SARS-Cov2-RT-28 | GCCATACTCCACTCATCTAAATC | SARS-Cov2-PCR-03 | GAAGAGATCGCCATTATTTTGGC |
| SARS-Cov2-RT-29 | CGCACAGAATTTTGAGCAGTTTC | SARS-Cov2-PCR-02N | AGAACTCGAAGGCATTCAGTAC |
| SARS-Cov2-RT-30 | CCCTCTTAGTGTCAATAAAGTCC | SARS-Cov2-PCR-01 | ATTAAAGGTTTATACCTTCCCAGGTAA |

**Patient sex and clade and lineage data for each of the 53 sequenced SARS-CoV-2 samples.**

| Patient sex | GISAID Clade | Pangolin Lineage | Sequencing technology |
|---|---|---|---|
| Female | S | A.2 | Illumina |
| Male | S | A.2 | Illumina |
| Female | S | A.2 | Illumina |
| Female | S | A.2 | Illumina |
| Male | S | A.2 | Illumina |
| Female | S | A.2 | Illumina |
| Female | S | A.2 | Illumina |
| Male | S | A.2 | Illumina |
| Female | S | A.2 | Illumina |
| Female | S | A.2 | Illumina |
| Male | S | A.2 | Illumina |
| Male | S | A.2 | Illumina |
| Male | S | A.2 | Illumina |
| Male | S | A.2 | Illumina |
| Female | S | A.2 | Illumina |
| Female | S | A.2 | Illumina |
| Female | S | A.2 | Illumina |
| unknown | S | A.2 | Sanger |
| Male | GH | B.1.255 | Illumina |
| Male | GH | B.1.255 | Illumina |
| unknown | G | B.1 | Sanger |
| Male | S | A.2 | Illumina |
| unknown | GR | B.1.1.10 | Sanger |
| Female | GH | B.1.3 | Illumina |
| Male | GH | B.1.3 | Illumina |
| Male | GH | B.1.3 | Illumina |
| Male | O | B.4.1 | Illumina |
| Male | O | B.4.1 | Illumina |
| Male | O | B.4.1 | Illumina |
| Female | O | B.4.1 | Illumina |
| Male | O | B.4.1 | Illumina |
| Female | O | B.4.1 | Illumina |
| Male | O | B.4.1 | Illumina |
| Female | O | B.4.1 | Illumina |
| Male | O | B.4.1 | Illumina |
| Male | O | B.4.1 | Illumina |
| Male | O | B.4.1 | Illumina |
| Male | O | B.4.1 | Illumina |
| Male | O | B.4.1 | Illumina |
| Male | O | B.4.1 | Illumina |
| Male | O | B.4.1 | Illumina |
| Male | O | B.4.1 | Illumina |
| Male | O | B.4.1 | Illumina |
| Male | O | B.4.1 | Illumina |
| Male | O | B.4.1 | Illumina |
| Male | O | B.4.1 | Illumina |
| Male | O | B.4.1 | Illumina |
| Male | O | B.4.1 | Illumina |
| Male | O | B.4.1 | Illumina |
| Male | O | B.4.1 | Illumina |
| Male | O | B.4.1 | Illumina |
| Male | O | B.4.1 | Illumina |
| unknown | O | B.4.1 | Sanger |

**Distribution of single-nucleotide polymorphisms (SNPs) in the Kazakhstan SARS-CoV-2 isolates (n=53) relative to the Wuhan-1 reference genome.**

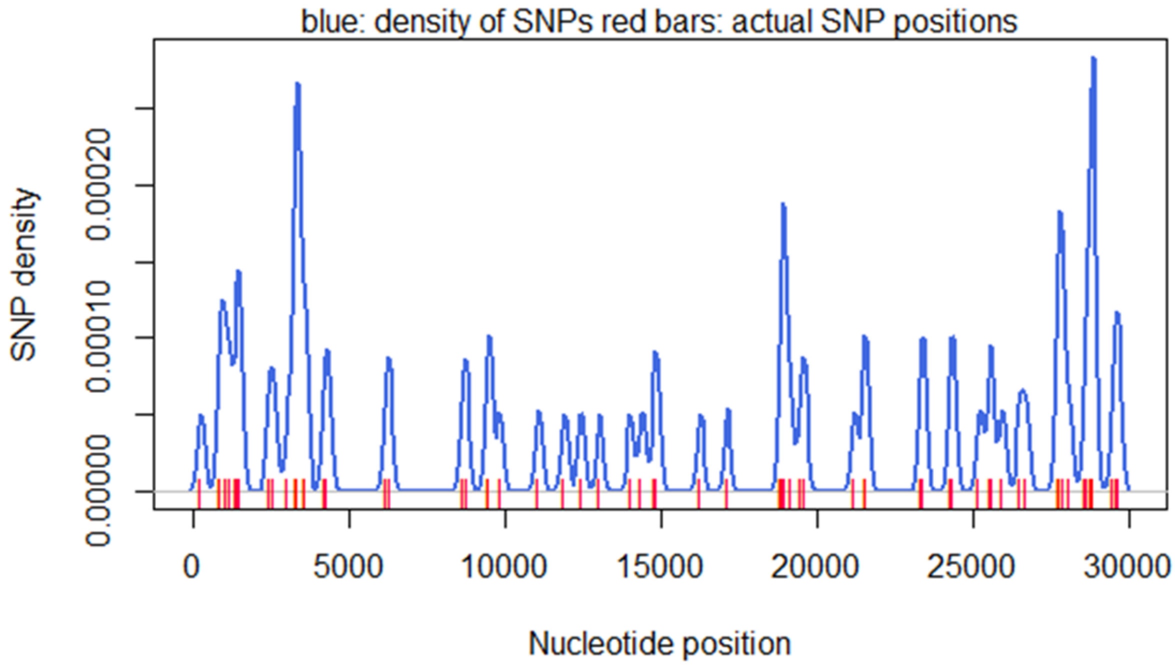

**Sample sizes of the demographic and clinical parameters used in the retrospective analysis**

| **Parameter** | **N of patients with available data** |
|---|---|
| Age | |
| Sex | 1072 |
| Ethnicity | |
| BMI | 93 |
| Days from symptom onset to admission | |
| Days in Hospital | 1072 |
| Comorbidities | |
| Body temperature | 779 |
| All other signs and symptoms | 1072 |
| Median pulse (absolute) | 764 |
| Pulse >125 beats/min (categorical) | 1072 |
| Median systolic pressure (absolute) | 686 |
| Syst pressure <90 mmHg (categorical) | 1072 |
| Median diastolic pressure (absolute) | 687 |
| Median respiratory rate (absolute) | 711 |
| Resp rate >24 breaths/min (categorical) | 1072 |
| Median SpO2 | 525 |
| White blood cells, median | 1018 |
| Neutrophil count, median | 483 |
| Lymphocyte count, median | 686 |
| NLR | 467 |
| Haemoglobin | 1018 |
| Monocytes | 550 |
| Eosinophils | 234 |
| Platelet countmedian | 955 |
| Prothrombin time | 363 |
| Fibrinogen | 383 |
| Albumin | 349 |
| Alanine aminotransferase | 800 |
| Aspartate aminotransferase | 655 |
| Total bilirubin | 719 |
| Direct bilirubin | 323 |
| Glucose | 667 |
| Blood urea nitrogen | 690 |
| Creatinine | 712 |
| C-reactive protein | 231 |
| Sodium | 144 |
| Potassium | 158 |
| Calcium | 118 |
| Radiologic findings | 737 |
| Treatments | 1072 |

**Members of the Semey COVID-19 Epidemiology Research Group (SCERG) and COVID-19 Genomics Research Group (CGRG) collaborative groups**

| **Members of the Semey COVID-19 Epidemiology Research Group (SCERG)** | |
|---|---|
| **Full Name** | **Affiliation** |
| Aidos Kusainov | Department of Surgical Disciplines, Semey Medical University, Semey, Kazakhstan |
| Aidana Rakhmankulova | Department of Clinical and Radiation Oncology, Semey Medical University, Semey, Kazakhstan |
| Aidos Tlemisov | Department of Surgery and Orthopedics, Semey Medical University, Semey, Kazakhstan |
| Aigul Kurmangazina | Department History of the Pediatric Surgery and Orthopedics, Semey Medical University, Semey, Kazakhstan |
| Aigul Omarova | Department of Biochemistry and Chemical Disciplines Semey Medical University, Semey, Kazakhstan |
| Ainash Orazalina | Department of Molecular Biology and Medical Genetics Semey Medical University, Semey, Kazakhstan |
| Alida Kaskabaeva | Department of Faculty Therapy, Semey Medical University, Semey, Kazakhstan |
| Aliya Alimbaeva | Department of Pediatrics, Semey Medical University, Semey, Kazakhstan |
| Alma Tokaeva | Department of Infectuous Diseases and Immunology, Semey Medical University, Semey, Kazakhstan |
| Almas Dyussupov | Department of Emergency Medicine, Semey Medical University, Semey, Kazakhstan |
| Almira Akhmetova | Department of Dermatological Venerology and Cosmetology, Semey Medical University, Semey, Kazakhstan |
| Altai Dyusupov | Department of Cardiovascular and Thoracic Surgery, Semey Medical University, Semey, Kazakhstan |
| Anastasia Griboyedova | Department of Pediatric Infectious diseases, Semey Medical University, Semey, Kazakhstan |
| Arailym Azharova | Department of Molecular Biology and Medical Genetics Semey Medical University, Semey, Kazakhstan |
| Ardak Zhumagaliyeva | Department of Hospital Therapy, Semey Medical University, Semey, Kazakhstan |
| Arna Abylkassymova | Department of Rheumatology and Non-Infectious Diseases, Semey Medical University, Semey, Kazakhstan |
| Askar Bukatov | Department of Topographical and Clinical Anatomy, Semey Medical University, Semey, Kazakhstan |
| Azhar Dyusupova | Department of Personified Medicine, Semey Medical University, Semey, Kazakhstan |
| Bakytkul Toktabayeva | Department of Propaedeutics of Childhood Diseases, Semey Medical University, Semey, Kazakhstan |
| Bakytzhan Alibekova | Department of Perinatology, Semey Medical University, Semey, Kazakhstan |
| Dariga Nurgaliyeva | Department of Hospital Therapy, Semey Medical University, Semey, Kazakhstan |
| Dariya Shabdarbayeva | Department Pathological anatomy and Forensic Medicine, Semey Medical University, Semey, Kazakhstan |
| Dinara Mukanova | Department of Simulation Technologies, Semey Medical University, Semey, Kazakhstan |
| Elmira Esimbekova | Department of Cardiology and Interventional Arrhythmology, Semey Medical University, Semey, Kazakhstan |
| Farida Rakhimzhanova | Department of Microbiology, Semey Medical University, Semey, Kazakhstan |
| Guliyash Tanysheva | Department of Obstetrics and Gynecology, Semey Medical University, Semey, Kazakhstan |
| Gulnar Berekenova | Department of Maxillofacial and Facial Plastic Surgery, Semey Medical University, Semey, Kazakhstan |
| Gulnara Shalgumbaeva | Department of Family Medicine, Semey Medical University, Semey, Kazakhstan |
| Gulnaz Kairatova | Department of Public Health and Evidence-Based Medicine, Semey Medical University, Semey, Kazakhstan |
| Gulshat Manabaeva | Department of Perinatology, Semey Medical University, Semey, Kazakhstan |
| Gulzhan Bersimbekova | Department of Rheumatology and Non-Infectious Diseases, Semey Medical University, Semey, Kazakhstan |
| Gulzhan Ilderbayeva | Department of Microbiology, Semey Medical University, Semey, Kazakhstan |
| Imdat Efendiev | Department of Pediatric infectious diseases, Semey Medical University, Semey, Kazakhstan |
| Kuralai Amrenova | Department of Propaedeutics of Internal Diseases, Semey Medical University, Semey, Kazakhstan |
| Laura Kasym | Department of Dermatological Venerology and Cosmetology, Semey Medical University, Semey, Kazakhstan |
| Madina Madiyeva | Department of Radiodiagnostic and Nuclear Medicine, Semey Medical University, Semey, Kazakhstan |
| Manar Salmenbayeva | Department of Pediatric infectious diseases, Semey Medical University, Semey, Kazakhstan |
| Marat Syzdykbaev | Department of Anesthesiology and Resuscitation Science, Semey Medical University, Semey, Kazakhstan |
| Maxut Kazymov | Department of Family Medicine, Semey Medical University, Semey, Kazakhstan |

| | |
|---|---|
| Meyerbek Aimagambetov | Department of Hospital Surgery, Semey Medical University, Semey, Kazakhstan |
| Muratkhan Kuderbaev | Department of Surgical Disciplines, Semey Medical University, Semey, Kazakhstan |
| Nurgul Barkibaeva | Department of Faculty Therapy, Semey Medical University, Semey, Kazakhstan |
| Nursultan Seksenbaev | Department of Psychiatry, Semey Medical University, Semey, Kazakhstan |
| Nurzhanat Haydarova | Department of Therapeutic Dentistry, Semey Medical University, Semey, Kazakhstan |
| Olga Gorkovenko | Department of Cardiovascular and Thoracic Surgery, Semey Medical University, Semey, Kazakhstan |
| Olga Van | Department of Topographical and Clinical Anatomy, Semey Medical University, Semey, Kazakhstan |
| Raushan Dinzhumanova | Department of Biochemistry and Chemical Disciplines, Semey Medical University, Semey, Kazakhstan |
| Raykhan Tuleutaeva | Department of Pharmacology named after Professor M. Mussin, Semey Medical University, Semey, Kazakhstan |
| Saltanat Uzbekova | Department Histology, Semey Medical University, Semey, Kazakhstan |
| Saule Kozhanova | Department Anatomy, Semey Medical University, Semey, Kazakhstan |
| Saule Maukaeva | Department of Infectuous Diseases and Immunology, Semey Medical University, Semey, Kazakhstan |
| Saule Rakhyzhanova | Department of Normal Physiology, Semey Medical University, Semey, Kazakhstan |
| Sholpan Abralina | Department of Pediatric Dentistry, Semey Medical University, Semey, Kazakhstan |
| Sholpan Zhukusheva | Department of Cardiology and Interventional Arrhythmology, Semey Medical University, Semey, Kazakhstan |
| Tasbolat Adylkhanov | Department of Clinical and Radiation Oncology, Semey Medical University, Semey, Kazakhstan |
| Timur Moldagaliev | Department of Psychiatry, Semey Medical University, Semey, Kazakhstan |
| Ulzhan Dzhamedinova | Department of Epidemiology and Biostatistics, Semey Medical University, Semey, Kazakhstan |
| Yulia Popovich | Department of Pediatrics, Semey Medical University, Semey, Kazakhstan |
| Yuliya Semenova | Department of Neurology and Neurophysiology, Semey Medical University, Semey, Kazakhstan |
| Zaytuna Hismetova | Department of Public Health and Evidence-Based Medicine, Semey Medical University, Semey, Kazakhstan |
| Zhanar Zamanbekova | Department of Endocrinology, Semey Medical University, Semey, Kazakhstan |
| Zhanar Zhumanbaeva | Department of Nursing, Semey Medical University, Semey, Kazakhstan |
| Zhanargul Smailova | Department of Biochemistry and Chemical Disciplines Named, Semey Medical University, Semey, Kazakhstan |
| Zhanna Kozykenova | Department of Pathological Physiology, Semey Medical University, Semey, Kazakhstan |
| **Members of the COVID-19 Genomics Research Group (CGRG) collaborative group** | |
| **Full Name** | **Affiliation** |
| Askar Abdaliyev | National Centre of Expertise, Nur Sultan, Kazakhstan |
| Baurzhan Negmetzhanov | Department of Biology, School of Science and Humanities, Nazarbayev University; National Laboratory Astana, Center for Life Sciences, Nazarbayev University, Nur-Sultan, Kazakhstan |
| Dinara Kamalova | National Centre for Biotechnology, Nur Sultan, Kazakhstan |
| Erlan Ramanculov | National Center for Biotechnology, Nur Sultan, Kazakhstan;<br>School of Science and Technology Nazarbayev University, Nur Sultan, Kazakhstan |
| Ilyas Akhmetollaev | National Center for Biotechnology, Nur Sultan, Kazakhstan |
| Kanat Balykbayev | National Centre of Expertise, Nur Sultan, Kazakhstan |
| Ruslan Kalendar | National Centre for Biotechnology, Nur Sultan, Kazakhstan |
| Sagyndyk Sagadinov | Chemistry-Biology Program, Suleyman Demirel University |
| Zabira Aushakhmetova | National Centre of Expertise, Nur Sultan, Kazakhstan |

**List of GISAID authors and laboratories, whose genomic data were used in the manuscript.**

| # | Authors | Strains | Laboratory |
|---|---|---|---|
| 1 | Leon Caly, Torsten Seemann, Michelle Sait, Mark B. Schultz, Julian Druce, Norelle L. Sherry | hCoV-19/Australia/VIC1566/2020 | Victorian Infectious Diseases Reference Laboratory (VIDRL) Microbiological Diagnostic Unit Public Health Laboratory and Victorian Infectious Diseases Reference Laboratory, Doherty Institute Caly L. et al https:www.gisaid.org NA NA 6/2/2020 |
| 2 | Alexandra Popa, Benedikt Agerer, Henrique Colaco, Lukas Endler, Jakob-Wendelin Genger, Alexander Lercher, Mark Smyth, Thomas Penz, Michael Schuster, Judith Aberle, Stephan Aberle, Elisabeth Puchhammer-Stцckl, Christoph Bock, Andreas Bergthaler | hCoV-19/Austria/CeMM0020/2020 : hCoV-19/Austria/CeMM0021/2020 | Center for Virology, Medical University of Vienna Bergthaler laboratory, CeMM Research Center for Molecular Medicine of the Austrian Academy of Sciences Alexandra Popa et al https:__www.gisaid.org Mutational dynamics and transmission properties of SARS-CoV-2 superspreading events in Austria https:__www.biorxiv.org_content_10.1101_2020.07.15.204339v1 4/3/2020 |
| 3 | Bert Vanmechelen, Joan Marti-Carreras, Tony Wawina, Piet Maes | hCoV-19/Belgium/RS-030257/2020 : hCoV-19/Belgium/VPE-030650/2020 : hCoV-19/Belgium/MMJ-03034/2020 | KU Leuven, Clinical and Epidemiological Virology KU Leuven, Clinical and Epidemiological Virology Bert Vanmechelen et al https:__www.gisaid.org A phylodynamic workflow to rapidly gain insights into the dispersal history and dynamics of SARS-CoV-2 lineages https:__dx.doi.org_10.1101_2020.05.05.078758 3/31/2020 |
| 4 | Durkin Keith, Artesi Maria, Bontems Sébastien, Boreux Raphaël, Meex Cécile, Melin Pierrette, Hayette Marie-Pierre, Bours Vincent. | hCoV-19/Belgium/ULG-6939/2020 | Department of Clinical Microbiology GIGA Medical Genomics Durkin Keith et al https:__www.gisaid.org A phylodynamic workflow to rapidly gain insights into the dispersal history and dynamics of SARS-CoV-2 lineages https:__dx.doi.org_10.1101_2020.05.05.078758 3/23/2020 |
| 5 | Qing Nie, Xingguang Li, Erik M Volz, Han Fu, Haowei Wang, Xiaoyue Xi, Wei Chen, Dehui Liu, Yingying Chen, Mengmeng Tian, Wei Tan, Junjie Zai, Wanying Sun, Jiandong Li, Junhua Li | hCoV-19/Weifang/WF0015/2020 | Weifang Center for Disease Control and Prevention Weifang Center for Disease Control and Prevention & BGI-Shenzhen Qing Nie et al https:__www.gisaid.org Genomic epidemiology of a densely sampled COVID19 outbreak in China https:__dx.doi.org_10.1101_2020.03.09.20033365 3/9/2020 |
| 6 | Filip Rokic, Lovro Trgovec-Greif, Neven Sucic, Tomislav Rukavina, Igor Jurak, Oliver Vugrek | hCoV-19/Croatia/LG-S2/2020 | Institute for Public Health Laboratory for advanced genomics Filip RokiÄ‡ et al https:__www.gisaid.org NA NA 4/24/2020 |
| 7 | Rasmus Kirkegaard | hCoV-19/Denmark/ALAB-HH81/2020 | Department of Clinical Microbiology, Copenhagen University Hospital, Hvidovre, Kettegaard Alle 30, 2650 Hvidovre. Albertsen lab, Department of Chemistry and Bioscience, Aalborg University, Denmark Rasmus Kirkegaard et al https:__www.gisaid.org NA NA 4/23/2020 |
| 8 | Monica Galiano, Shahjahan Miah, Angie Lackenby, Omolola Akinbami, Tiina Talts, Leena Bhaw, Richard Myers, Steven Platt, Kirstin Edwards, Jonathan Hubb, Joanna Ellis, Maria Zambon | hCoV-19/England/01/2020 : hCoV-19/England/200990724/2020 : hCoV-19/England/20102000306/2020 : hCoV-19/England/20109053606/2020 : hCoV-19/England/20126006006/2020 : hCoV-19/England/20126027006/2020 : hCoV-19/England/20129061704/2020 : hCoV-19/England/20134019904/2020 : hCoV-19/England/20134021604/2020 : hCoV- | Respiratory Virus Unit, Microbiology Services Colindale, Public Health England Respiratory Virus Unit, Microbiology Services Colindale, Public Health England Monica Galiano et al https:__www.gisaid.org Preliminary analysis of SARS-CoV-2 importation & establishment of UK transmission lineages https:__virological.org_t_preliminary-analysis-of-sars-cov-2-importation-establishment-of-uk-transmission-lineages_507 3/10/2020 |

| | | | |
|---|---|---|---|
| | | 19/England/201 39018302/2020 | |
| 9 | William J. Liu, Peipei Liu, Xiang Zhao, Peihua Niu, Yingze Zhao, Wenwen Lei, Ziqian Xu, Shumei Zou, Wei Zhen, Beiwei Ye, Mengjie Yang, Weifeng Shi, Roujian Lu, Wenjie Tan, Zhixiao Chen, Yuchao Wu, Juan Song, Weimin Zhou, Dayan Wang, Jun Han, Wenbo Xu, George F. Gao, Guizhen Wu | hCoV-19/env/Wuhan/IVDC-HBF13-20/2020 : hCoV-19/env/Wuhan/IVDC-HBF13-21/2020 | Institute of Viral Disease Control and Prevention, China CDC Institute of Viral Disease Control and Prevention, China CDC William J. Liu et al https:__www.gisaid.org NA NA 2/9/2020 |
| 10 | Teemu Smura, Hannimari Kallio-Kokko, Olli Vapalahti | hCoV-19/Finland/13M57/2020 | Department of Virology and Immunology, University of Helsinki and Helsinki University Hospital, Huslab Finland Department of Virology, Faculty of Medicine, University of Helsinki, Helsinki, Finland Teemu Smura et al https:__www.gisaid.org NA NA 3/30/2020 |
| 11 | Rodriguez,C., De Prost,N., Fourati,S., Lamoureux,C., Schmitz,D., Deveaux,I., Picard,O., Lepeule,R., Surgers,L., Mekontso-Dessap,A., Woerther,P.-L., Canoui-Poitrine,F., Pawlotsky,J.-M., Clinical Study Group,C., Gricourt,G., N'debi,M., Demontant,V., Trawinski,E. | hCoV-19/France/IDF-10062MD/2020 | Hôpital Henri-Mondor Ap-Hpn Genomic platform De Prost et al https:__www.gisaid.org NA NA 5/14/2020 |
| 12 | Mélnie Albert, Marion Barbet, Sylvie Behillil, Méline Bizard, Angela Brisebarre, Flora Donati, Etienne Simon-Lorière, Vincent Enouf, Maud Vanpeene, Sylvie van der Werf, Gisèle Lagathu | hCoV-19/France/BRE-2348/2020 : hCoV-19/France/GES-1583/2020 : hCoV-19/France/HDF-3141/2020 : hCoV-19/France/IDF-0372-isl/2020 : hCoV-19/France/IDF-0515-isl/2020 : hCoV-19/France/IDF-2561/2020 | CHRU Pontchaillou - Laboratoire de Virologie National Reference Center for Viruses of Respiratory Infections, Institut Pasteur, Paris MÃ©lnie Albert et al https:__www.gisaid.org Introductions and early spread of SARS-CoV-2 in France https:__dx.doi.org_10.1101_2020.04.24.059576 3/22/2020 |
| 13 | Marine Murtskhvaladze, Nato Kotaria, Ann Machablishvili, Lela Sabadze, Mari Gavashelidze, Ana Papkiauri, Meri Pantsulaia, Gvantsa Brachveli, Tata Imnadze, Tamar Jashiashvili, Tea Tevdoradze, Ketevan Sidamonidze, Ekaterine Khmaladze, Ekaterine Zhghenti, Roena Sukhiashvili, Mariam Zakalashvili, Lela Urushadze, Magda Dgebuadze, Giorgi Tomashvili, Davit Tsaguria, Ekaterine Zangaladze, Nino Berishvili, Gvantsa Chanturia, Adam Kotorashvili, Maia Alkhazashvili, Irma Burjanadze, Anna Kasradze, Khatuna Zakhashvili, Paata Imnadze, Amiran Gamkrelidze. | hCoV-19/Georgia/Tb-390/2020 | R. G. Lugar Center for Public Health Research, National Center for Disease Control and Public Health (NCDC) of Georgia. R. G. Lugar Center for Public Health Research, National Center for Disease Control and Public Health (NCDC) of Georgia. Marine Murtskhvaladze et al https:__www.gisaid.org NA NA 3/22/2020 |
| 14 | Victor M Corman, Julia Schneider, Barbara Mühlemann, Talitha Veith, Jörn Beheim-Schwarzbach, Terry Jones, Rainer Oehme, Silke Fischer, Christian Drosten | hCoV-19/Germany/BW-ChVir-1577/2020 | State Health Office Baden-Wuerttemberg CharitÃ© UniversitÃ¤tsmedizin Berlin, Institute of Virology Victor M Corman et al https:__www.gisaid.org Whole genome and phylogenetic analysis of two SARS-CoV-2 strains isolated in Italy in January and February 2020:00:00 additional clues on multiple introductions and further circulation in Europe https:__dx.doi.org_10.2807_1560-7917.ES.2020.25.13.2000305 2/28/2020 |

| | | | |
|---|---|---|---|
| 15 | Boehmer,M.M., Buchholz,U., Corman,V.M., Hoch,M., Katz,K., Marosevic,D.V., Boehm,S., Woudenberg,T., Ackermann,N., Konrad,R., Eberle,U., Treis,B., Dangel,A., Bengs,K., Fingerle,V., Berger,A., Hoermansdorfer,S., Ippisch,S., Wicklein,B., Grahl,A., Poertner,K., Muller,N., Zeitlmann,N., Boender,T.S., Cai,W., Reich,A., an der Heiden,M., Rexroth,U., Hamouda,O., Schneider,J., Veith,T., Muehlemann,B., Woelfel,R., Antwerpen,M., Walter,M., Protzer,U., Liebl,B., Haas,W., Sing,A., Drosten,C., Zapf,A., Jones,T.C. | hCoV-19/Germany/BY-ChVir-1459/2020 | unknown Department of Virology Boehmer et al https:__www.gisaid.org NA NA 5/19/2020 |
| 16 | Huang,J., Pfefferle,S. and Fischer,N. | hCoV-19/Germany/HH-UMC-01/2020 | unknown Center for Diagnostics Huang et al https:__www.gisaid.org Complete Genome Sequence of a SARS-CoV-2 Strain Isolated in Northern Germany https:__dx.doi.org_10.1128_MRA.00520-20 5/4/2020 |
| 17 | Ortwin Adams, Marcel Andree, Alexander Dilthey, Torsten Feldt, Sandra Hauka, Torsten Houwaart, Björn-Erik Jensen, Detlef Kindgen-Milles, Malte Kohns Vasconcelos, Klaus Pfeffer, Tina Senff, Daniel Strelow, Jörg Timm, Andreas Walker, Tobias Wienemann | hCoV-19/Germany/NW-HHU-31/2020 | Center of Medical Microbiology, Virology, and Hospital Hygiene, University of Duesseldorf Center of Medical Microbiology, Virology, and Hospital Hygiene, University of Duesseldorf Ortwin Adams et al https:__www.gisaid.org Genetic structure of SARS-CoV-2 in Western Germany reflects clonal superspreading and multiple independent introduction events https:__dx.doi.org_10.1101_2020.04.25.20079517 4/2/2020 |
| 18 | Kassela K., Bampali,M., Dovrolis,N., Gatzidou,E., Froukala,E., Stavropoulou,A., Veletza,S., Tsakris,A., Spanakis,N. and Karakasiliotis,I. | hCoV-19/Greece/136/2020 | Laboratory of Microbiology, Medical School, National and Kapodistrian University of Athens Laboratory of Biology, Department of Medicine, Democritus University of Thrace Kassela K. et al https:__www.gisaid.org Dominant and rare SARS-Cov2 variants responsible for the COVID-19 pandemic in Athens, Greece. https:__dx.doi.org_10.1101_2020.06.03.20121236 4/29/2020 |
| 19 | Jing Lu, Louis du Plessis, Liu Zhe, Jiufeng Sun, Sarah François, Huifang Lin, Moritz Kraemer, Jingju Peng, Qianlin Xiong, Runyu Yuan, Lilian Zeng, Pingping Zhou, Chuming Liang, Tao Liu, Wei Li, Juan Su, Huanying Zheng, Kang Min, Song Tie, Bo Peng, Shisong Fang, Wenzhe Su, Kuibiao Li, Ruilin Sun, Ru bai, Xi Tang, Minfeng Liang, Nuno Faria, Josh Quick, Andrew Rambaut, Verity Hill, Wenjun Ma, Nick Loman, Oliver Pybus, Changwen Ke | hCoV-19/Guangdong/2020XN4475-P0042/2020 : hCoV-19/Guangdong/GD2020087-P0008/2020 | Guangdong Provincial Institution of Public Health, Guangdong Provinical Center for Disease Control and Prevention Guangdong Provincial Institution of Public Health Jing Lu et al https:__www.gisaid.org Genomic Epidemiology of SARS-CoV-2 in Guangdong Province, China https:__dx.doi.org_10.1016_j.cell.2020.04.023 3/9/2020 |
| 20 | Bosheng Li, Haogao Gu, Lijun Liang, Zhencui Li, Hui-Ling Yen, Yao Hu, Yingchao Song , Hanri Zeng, Tie Song, Jie Wu, Leo L.M. Poon | hCoV-19/Guangdong/20SF198/2020 | Guangdong Provincial Center for Diseases Control and Prevention;Guangdong Provincial Institute of Public Health School of Public Health, The University of Hong Kong Bosheng Li et al https:__www.gisaid.org NA NA 4/21/2020 |
| 21 | Li,X., Lu,S., Wu,B., Hu,X., Li,D., Huang,X. and Guo,W. | hCoV-19/Henan/HN03/2020 | unknown The Department of Infectious Disease Prevention and Control Li et al https:__www.gisaid.org NA NA 4/30/2020 |
| 22 | Lau,S.K.P., Luk,H.K.H., Wong,A.C.P., Li,K.S.M., Zhu,L., He,Z., Fung,J., Chan,T.T.Y., Fung,K.S.C. and Woo,P.C.Y. | hCoV-19/Hong Kong/HK20/2020 | unknown Department of Microbiology Lau et al https:__www.gisaid.org NA NA 4/24/2020 |
| 23 | Péter Urbán, Endre Gábor Tóth, Gábor Kemenesi, Róbert Herczeg, Attila Gyenesei, Ferenc Jakab | hCoV-19/Hungary/mbl49/2020 | Virological Research Group, SzentÃ¡gothai Research Centre Bioinformatics Research Group, SzentÃ¡gothai Research Centre PÃ©ter UrbÃ¡n et al https:__www.gisaid.org NA NA 3/23/2020 |
| 24 | Daniel F Gudbjartsson; Agnar Helgason; Hakon Jonsson; Olafur T Magnusson; Pall Melsted; Gudmundur L Norddahl; Jona Saemundsdottir; Asgeir Sigurdsson; Patrick Sulem; Arna B Agustsdottir; Berglind Eiriksdottir; Run Fridriksdottir; Elisabet E Gardarsdottir; Gudmundur Georgsson; Olafia S Gretarsdottir; Kjartan R Gudmundsson; Thora R Gunnarsdottir; Arnaldur Gylfason; Hilma Holm; Brynjar O Jensson; Aslaug Jonasdottir; Kamilla S Josefsdottir; Thordur Kristjansson; Droplaug N Magnusdottir; Louise le Roux; Gudrun Sigmundsdottir; Gardar | hCoV-19/Iceland/259/2020 | The National University Hospital of Iceland deCODE genetics Daniel F Gudbjartsson et al https:__www.gisaid.org Spread of SARS-CoV-2 in the Icelandic Population https:__dx.doi.org_10.1056_NEJMoa2006100 3/27/2020 |

| | | | |
|---|---|---|---|
| | Sveinbjornsson; Kristin E Sveinsdottir; Maney Sveinsdottir; Emil A Thorarensen; Bjarni Thorbjornsson; Gisli Masson; Ingileif Jonsdottir; Alma Moller; Thorolfur Gudnason; Karl G Kristinsson; Unnur Thorsteinsdottir; Kari Stefansson | | |
| 25 | Khosravi,M.A., Abbasalipour,M., Zeinali,S., Sabeghi,S., Kehsvar,Y., Hosseini,F. and Haghdoost,Y. | hCoV-19/Iran/HGRC-2-2162/2020 | unknown Human Genetic Research Center Khosravi et al https:__www.gisaid.org NA NA 5/8/2020 |
| 26 | Sirous Zeinali, Mohammad Ali Khosravi,Maryam Abbasalipour Bashash, Sanaz Mostafavi Jabbari, Maraym Firoozi, Sormeh Pourtavakoli, Elmira Khateri, Razieh Zeinali and Fahimeh Hoseini | hCoV-19/Iran/KHGRC-1.1-IPI-8206/2020 | Pasteur Institute of Iran Kawsar Human Genetic Research Company Sirous Zeinali et al https:__www.gisaid.org NA NA 5/12/2020 |
| 27 | Mohammad Ali Khosravi, Maryam Abbasalipour Bashash, Sirous Zeinali, Solmaz Sabeghi, Yeganeh Keshvar, Fatemeh Hosseini, Yeganeh Haghdoost | hCoV-19/Iran/KHGRC-2-2162/2020 | Kawsar Human Genetic Research Center Kawsar Human Genetic Research Center Mohammad Ali Khosravi et al https:__www.gisaid.org NA NA 5/12/2020 |
| 28 | Paola Stefanelli, Stefano Fiore, Antonella Marchi, Eleonora Benedetti, Concetta Fabiani, Giovanni Faggioni, Antonella Fortunato, Riccardo De Santis, Silvia Fillo, Anna Anselmo, Andrea Ciammaruconi, Stefano Palomba, Florigio Lista | hCoV-19/Italy/LOM-ASST-CDG1/2020 | Department of Infectious Diseases, Istituto Superiore di SanitÃ , Roma , Italy Virology Laboratory, Scientific Department, Army Medical Center Paola Stefanelli et al https:__www.gisaid.org Whole genome and phylogenetic analysis of two SARS-CoV-2 strains isolated in Italy in January and February 2020:00:00 additional clues on multiple introductions and further circulation in Europe https:__dx.doi.org_10.2807_1560-7917.ES.2020.25.13.2000305 3/1/2020 |
| 29 | Maria R. Capobianchi, Cesare E. M. Gruber, Martina Rueca, Barbara Bartolini, Francesco Messina, Emanuela Giombini, Francesca Colavita, Concetta Castilletti, Eleonora Lalle, Fabrizio Carletti, Emanuele Nicastri, Giuseppe Ippolito. | hCoV-19/Italy/LAZ-INMI1-isl/2020 : hCoV-19/Italy/LAZ-INMI3/2020 : hCoV-19/Italy/LAZ-INMI4/2020 | INMI Lazzaro Spallanzani IRCCS Laboratory of Virology, INMI Lazzaro Spallanzani IRCCS Maria R. Capobianchi et al https:__www.gisaid.org Whole genome and phylogenetic analysis of two SARS-CoV-2 strains isolated in Italy in January and February 2020:00:00 additional clues on multiple introductions and further circulation in Europe https:__dx.doi.org_10.2807_1560-7917.ES.2020.25.13.2000305 2/17/2020 |
| 30 | Paola Stefanelli, Stefano Fiore, Antonella Marchi, Eleonora Benedetti, Concetta Fabiani, Giovanni Faggioni, Antonella Fortunato, Silvia Fillo, Riccardo De Santis, Andrea Ciammaruconi, Giancarlo Petralito, Filippo Molinari, Florigio Lista | hCoV-19/Italy/LAZ-INMI-SPL1/2020 : hCoV-19/Italy/LAZ-INMI-SPL1/2020 | Department of Infectious Diseases, Istituto Superiore di SanitÃ , Rome, Italy Virology Laboratory, Scientific Department, Army Medical Center Paola Stefanelli et al https:__www.gisaid.org Whole genome and phylogenetic analysis of two SARS-CoV-2 strains isolated in Italy in January and February 2020:00:00 additional clues on multiple introductions and further circulation in Europe https:__dx.doi.org_10.2807_1560-7917.ES.2020.25.13.2000305 3/1/2020 |

| | | | |
|---|---|---|---|
| 31 | Lorusso A, Marcacci M, Di Domenico M, Ancora M, Curini V, Mangone I, Rinaldi A, Di Pasquale A, Camma C, Puglia I, Savini G | hCoV-19/Italy/ABR-IZSGC-TE13457/2020 : hCoV-19/Italy/ABR-IZSGC-TE13858/2020 : hCoV-19/Italy/ABR-IZSGC-TE26423/2020 : hCoV-19/Italy/ABR-IZSGC-TE26425/2020 : hCoV-19/Italy/ABR-IZSGC-TE5545/2020 : hCoV-19/Italy/ABR-IZSGC-TE5551/2020 : hCoV-19/Italy/ABR-IZSGC-TE6225/2020 : hCoV-19/Italy/ABR-IZSGC-TE6644/2020 : hCoV-19/Italy/ABR-AMVRC-TE7859/2020 | Villa Serena del Dr. Leonardo Petruzzi Istituto Zooprofilattico Sperimentale dell?˘™Abruzzo e Molise ?ˆ?G. Caporale?ˆ• Lorusso A et al https:__www.gisaid.org NA NA 5/3/2020 : SERVIZIO DI IGIENE E SANIT?ˆ PUBBLICA ASL Teramo Istituto Zooprofilattico Sperimentale dell?˘™Abruzzo e Molise ?ˆ?G. Caporale?ˆ• Lorusso A et al https:__www.gisaid.org NA NA 5/3/2020 : Servizio di Igiene, Epidemiologia e Sanit? Pubblica (SIESP) Avezzano Istituto Zooprofilattico Sperimentale dell?˘™Abruzzo e Molise ?ˆ?G. Caporale?ˆ• Lorusso A et al https:__www.gisaid.org NA NA 5/3/2020 : Ospedale Civile S. Liberatore di Atri Istituto Zooprofilattico Sperimentale dell?˘™Abruzzo e Molise ?ˆ?G. Caporale?ˆ• Lorusso A et al https:__www.gisaid.org NA NA 5/7/2020 : Ospedale Civile S. Liberatore di Atri Istituto Zooprofilattico Sperimentale dell?˘™Abruzzo e Molise ?ˆ?G. Caporale?ˆ• Lorusso A et al https:__www.gisaid.org NA NA 5/7/2020 : Ospedale Civile S. Liberatore di Atri Istituto Zooprofilattico Sperimentale dell?˘™Abruzzo e Molise ?ˆ?G. Caporale?ˆ• Lorusso A et al https:__www.gisaid.org NA NA 5/7/2020 : Presidio Ospedaliero Santo Spirito Istituto Zooprofilattico Sperimentale dellAbruzzo e Molise "G. Caporale" Lorusso A et al https:__www.gisaid.org NA NA 4/23/2020 Ospedale Civile Giuseppe Mazzini Istituto Zooprofilattico Sperimentale dell?˘™Abruzzo e Molise ?ˆ?G. Caporale?ˆ• Lorusso A et al https:__www.gisaid.org NA NA 4/6/2020 Ospedale Civile Giuseppe Mazzini Istituto Zooprofilattico Sperimentale dell?˘™Abruzzo e Molise ?ˆ?G. Caporale?ˆ• Lorusso A et al https:__www.gisaid.org NA NA 5/3/2020 Ospedale Civile S. Liberatore di Atri Istituto Zooprofilattico Sperimentale dellAbruzzo e Molise G. Caporale Lorusso A et al https:__www.gisaid.org NA NA 4/23/2020 |
| 32 | Gianguglielmo Zehender, Alessia Lai, Annalisa Bergna, Luca Meroni, Agostino Riva, Claudia Balotta, Maciej Tarkowski, Arianna Gabrieli, Dario Bernacchia, Stefano Rusconi, Giuliano Rizzardini, Spinello Antinori, Massimo Galli | hCoV-19/Italy/LOM-UniMI01/2020 | Laboratory of Infectious Diseases, Department of Biomedical and Clinical Sciences L. Sacco, University of Milan Laboratory of Infectious Diseases, Department of Biomedical and Clinical Sciences L. Sacco, University of Milan Gianguglielmo Zehender et al https:__www.gisaid.org Genomic characterization and phylogenetic analysis of SARS-COV-2 in Italy https:__onlinelibrary.wiley.com_doi_abs_10.1002_jmv.25794 3/26/2020 |
| 33 | R.A Diotti, E. Criscuolo, M. Castelli, V. Caputo, R. Ferrarese, M. Sampaolo, E. Boeri, I. Negri, V. Amato, G. Lo Raso, C. Di Resta, R. Burioni, M. Clementi, N. Mancini & N. Clementi | hCoV-19/Italy/LOM-UniSR-1/2020 : hCoV-19/Italy/LOM-UniSR-1/2020 | Laboratorio di Microbiologia e Virologia, UniversitÃ Vita-Salute San Raffaele, Milano Laboratorio di Microbiologia e Virologia, UniversitÃ Vita-Salute San Raffaele, Milano R.A Diotti et al https:__www.gisaid.org NA NA 3/5/2020 |
| 34 | Kenjiro Kosaki, Yuka Iwasaki, Toshiki Takenouchi, Haruhiko Siomi | hCoV-19/Japan/Donner1/2020 : hCoV-19/Japan/Donner12/2020 : hCoV-19/Japan/Donner17/2020 : hCoV-19/Japan/Donner18/2020 : hCoV-19/Japan/Donner2/2020 : hCoV-19/Japan/Donner22/2020 : hCoV-19/Japan/Donner27/2020 : | Keio University School of Medicine Keio University School of Medicine Kenjiro Kosaki et al https:__www.gisaid.org Clinical Utility of SARS-CoV-2 Whole Genome Sequencing in Deciphering Source of Infection https:__dx.doi.org_10.1101_2020.05.21.20107599 5/11/2020 |

| | | | |
|---|---|---|---|
| | | hCoV-19/Japan/Donner28/2020 | |
| 35 | Tsuyoshi Sekizuka, Kentaro Itokawa, Rina Tanaka, Masanori Hashino, Tsutomu Kageyama, Shinji Saito, Ikuyo Takayama, Hideki Hasegawa, Takuri Takahashi, Hajime Kamiya, Takuya Yamagishi, Motoi Suzuki, Takaji Wakita, Makoto Kuroda | hCoV-19/Japan/DP0703/2020 : hCoV-19/Japan/DP0724/2020 | Japanese Quarantine Stations Pathogen Genomics Center, National Institute of Infectious Diseases Tsuyoshi Sekizuka et al https:__www.gisaid.org Haplotype networks of SARS-CoV-2 infections in the Diamond Princess cruise ship outbreak https:__dx.doi.org_10.1101_2020.03.23.20041970 3/23/2020 |
| 36 | Hishiki,T., Suzuki,R., Sakuragi,J., Usui,K., Tanaka,Y., Kawai,J., Kogo,Y., Matsuki,Y., An,T., Hayashizaki,Y. and Takasaki,T. | hCoV-19/Japan/Hu_DP_Kng_19-027/2020 : hCoV-19/Japan/Hu_DP_Kng_19-031/2020 : hCoV-19/Japan/Hu_Kng_19-437/2020 | unknown Takayuki Hishiki Kanagawa Prefectural Institute of Public Health, Department of Microbiology Hishiki et al https:__www.gisaid.org A doubt of multiple introduction of SARS-CoV-2 in Italy: A preliminary overview https:__dx.doi.org_10.1002_jmv.25773 2/29/2020 |
| 37 | Tsuyoshi Sekizuka, Shutoku Matsuyama, Naganori Nao, Kazuya Shirato, Makoto Takeda, Makoto Kuroda | hCoV-19/Japan/KY-V-029/2020 : hCoV-19/Japan/TY-WK-012/2020 : hCoV-19/Japan/TY-WK-521/2020 | Dept. of Virology III, National Institute of Infectious Diseases Pathogen Genomics Center, National Institute of Infectious Diseases Tsuyoshi Sekizuka et al https:__www.gisaid.org NA NA 2/10/2020 |
| 38 | Tsuyoshi Sekizuka, Michiyo Shinohara, Tsuyoshi Kishimoto, Kentaro Itokawa, Rina Tanaka, Masanori Hashino, Hajime Kamiya, Motoi Suzuki, Makoto Kuroda | hCoV-19/Japan/P4-7/2020 : hCoV-19/Japan/P4-8/2020 | Saitama Prefectural Institute of Public Health Pathogen Genomics Center, National Institute of Infectious Diseases Tsuyoshi Sekizuka et al https:__www.gisaid.org SARS-CoV-2 Genome Analysis of Japanese Travelers in Nile River Cruise https:__dx.doi.org_10.3389_fmicb.2020.01316 4/2/2020 |
| 39 | Tsuyoshi Sekizuka, Masakatsu Taira, Yushi Hachisu, Kentaro Itokawa, Rina Tanaka, Masanori Hashino, Hajime Kamiya, Motoi Suzuki, Makoto Kuroda | hCoV-19/Japan/P5-1/2020 : hCoV-19/Japan/P5-3/2020 | Chiba Prefectural Institute of Public Health Pathogen Genomics Center, National Institute of Infectious Diseases Tsuyoshi Sekizuka et al https:__www.gisaid.org SARS-CoV-2 Genome Analysis of Japanese Travelers in Nile River Cruise https:__dx.doi.org_10.3389_fmicb.2020.01316 4/2/2020 |
| 40 | Kazuo Imai | hCoV-19/Japan/SMU-0311S3/2020 | Saitama Medical University Saitama Medical University Kazuo Imai et al https:__www.gisaid.org NA NA 3/22/2020 |
| 41 | Kumagai,R., Yoshida,I., Asakura,H., Nagashima,M., Chiba,T. and Sadamasu,K. | hCoV-19/Japan/TKYE6182/2020 : hCoV-19/Japan/TKYE6947/2020 : hCoV-19/Japan/TKYE6968/2020 | unknown Ryota Kumagai Tokyo Metropolitan Institute of Public Health Kumagai et al https:__www.gisaid.org NA NA 3/13/2020 |
| 42 | Bin Fang, Xiang Li, Xiao Yu, Linlin Liu, Bo Yang, Faxian Zhan, Guojun Ye, Xixiang Huo, Junqiang Xu, Bo Yu, Kun Cai, Jing Li, Maoyi Chen,Jie Hu, Chunlin Mao, Yongzhong Jiang. | hCoV-19/Jingzhou/HBCDC-HB-01/2020 | Jingzhou Center for Disease Control and Prevention Hubei Provincial Center for Disease Control and Prevention Bin Fang et al https:__www.gisaid.org Genome-wide data inferring the evolution and population demography of the novel pneumonia coronavirus (SARS-CoV-2) https:__dx.doi.org_10.1101_2020.03.04.976662 2/26/2020 |

| | | | |
|---|---|---|---|
| 43 | Alexandr Shevtsov, Ilyas Akhmetollayev, Viktoriya Lutsay, Asylulan Amirgazin, Askar Abdaliyev, Akbota Rakhmetova, Zabira Aushakhmetova, Ruslan Kalendar, Yerlan Ramankulov | hCoV-19/Kazakhstan/16173/2020 : hCoV-19/Kazakhstan/16183/2020 : hCoV-19/Kazakhstan/16238/2020 : hCoV-19/Kazakhstan/16537/2020 : hCoV-19/Kazakhstan/17920/2020 : hCoV-19/Kazakhstan/18148/2020 : hCoV-19/Kazakhstan/18243/2020 : hCoV-19/Kazakhstan/18287/2020 : hCoV-19/Kazakhstan/20679/2020 : hCoV-19/Kazakhstan/21399/2020 : hCoV-19/Kazakhstan/21927/2020 : hCoV-19/Kazakhstan/22001/2020 : hCoV-19/Kazakhstan/22044/2020 : hCoV-19/Kazakhstan/22517/2020 : hCoV-19/Kazakhstan/26473/2020 : hCoV-19/Kazakhstan/26474/2020 : hCoV-19/Kazakhstan/26474/2020 : hCoV-19/Kazakhstan/26478/2020 : hCoV-19/Kazakhstan/26489/2020 : hCoV-19/Kazakhstan/26491/2020 : hCoV-19/Kazakhstan/26497/2020 : hCoV-19/Kazakhstan/26501/2020 : hCoV-19/Kazakhstan/26506/2020 : hCoV- | Laboratory of virology, National Center of Expertise Laboratory of molecular-genetic research, National Center of Expertise, Kazakhstan National Center for Biotechnology, Kazakhstan Abdaliyev Askar et al https__www.gisaid.org NA NA 5/28/2020 |

| | | |
|---|---|---|
| | | 19/Kazakhstan/26508/2020 : hCoV-19/Kazakhstan/26530/2020 : hCoV-19/Kazakhstan/26545/2020 : hCoV-19/Kazakhstan/26548/2020 : hCoV-19/Kazakhstan/26549/2020 : hCoV-19/Kazakhstan/26562/2020 : hCoV-19/Kazakhstan/26565/2020 : hCoV-19/Kazakhstan/26568/2020 : hCoV-19/Kazakhstan/26574/2020 : hCoV-19/Kazakhstan/26576/2020 : hCoV-19/Kazakhstan/26577/2020 : hCoV-19/Kazakhstan/26579/2020 : hCoV-19/Kazakhstan/26580/2020 : hCoV-19/Kazakhstan/26582/2020 : hCoV-19/Kazakhstan/26584/2020 : hCoV-19/Kazakhstan/26585/2020 : hCoV-19/Kazakhstan/26617/2020 : hCoV-19/Kazakhstan/26827/2020 : hCoV-19/Kazakhstan/26828/2020 : hCoV-19/Kazakhstan/26829/2020 : hCoV-19/Kazakhstan/33496/2020 : hCoV-19/Kazakhstan/34391/2020 : hCoV-19/Kazakhstan/38533/2020 : hCoV-19/Kazakhstan/3 |

| | | | |
|---|---|---|---|
| | | 8716/2020 : hCoV-19/Kazakhstan/7263/2020 : hCoV-19/Kazakhstan/7341/2020 : hCoV-19/Kazakhstan/NCB-1/2020 : hCoV-19/Kazakhstan/NCB-2/2020 : hCoV-19/Kazakhstan/NCB-3/2020 : hCoV-19/Kazakhstan/NCB-5/2020 | |
| 44 | Krista Queen, Yan Li, Ying Tao, Jing Zhang, Anne Uehara, Clinton R. Paden, Haibin Wang, Rachel Marine, Mary S. Keckler, Alison S. Laufer Halpin, Jasmine Padilla, Justin Lee, Christopher A. Elkins, Suxiang Tong | hCoV-19/South Korea/BA-ACH_2718/2020 | Brian D. Allgood Army Community Hospital Pathogen Discovery, Respiratory Viruses Branch, Division of Viral Diseases, Centers for Disease Control and Prevention Krista Queen et al https:__www.gisaid.org NA NA 4/6/2020 |
| 45 | Jeong-Min Kim, Yoon-Seok Chung, Namjoo Lee, Mi-Seon Kim, Sang Hee Woo, Hye-Jun Jo, Sehee Park, Heui Man Kim, Jun-Sub Kim, Junhyeong Jang, Dong Hyun Song, Daesang Lee, Seong Tae Jeong, Myung Guk Han | hCoV-19/South Korea/KCDC2001/2020 : hCoV-19/South Korea/KCDC2002/2020 : hCoV-19/South Korea/KCDC2005/2020 : hCoV-19/South Korea/KCDC2008/2020 : hCoV-19/South Korea/KCDC2009/2020 : hCoV-19/South Korea/KCDC2011/2020 : hCoV-19/South Korea/KCDC2014/2020 : hCoV-19/South Korea/KCDC2019/2020 | Division of Viral Diseases, Center for Laboratory Control of Infectious Diseases, Korea Centers for Diseases Control and Prevention Division of Viral Diseases, Center for Laboratory Control of Infectious Diseases, Korea Centers for Diseases Control and Prevention Jeong-Min Kim et al https:__www.gisaid.org NA NA 4/14/2020 |
| 46 | Jeong-Min Kim, Yoon-Seok Chung, Namjoo Lee, Mi-Seon Kim, SangHee Woo, Hye-Joon Jo, Sehee Park, Heui Man Kim, Myung Guk Han | hCoV-19/South Korea/KCDC03/2020 | Korea Centers for Disease Control & Prevention (KCDC) Center for Laboratory Control of Infectious Diseases Division of Viral Diseases Korea Centers for Disease Control & Prevention (KCDC) Center for Laboratory Control of Infectious Diseases Division of Viral Diseases Jeong-Min Kim et al https:__www.gisaid.org Identification of Coronavirus Isolated from a Patient in Korea with COVID-19 https:__www.ncbi.nlm.nih.gov_pmc_articles_PMC7045880_ 2/4/2020 |
| 47 | Fahd Al-Mulla, Sumi John, Rasheeba Iqbal, Motasem Melhem, Ebaa AlOzairi, Sara Al-Qabandi, Qais Al-Duwairi | hCoV-19/Kuwait/KU17/2020 | Dasman Diabetes Institute Dasman Diabetes Institute Fahd Al-Mulla et al https:__www.gisaid.org NA NA 3/23/2020 |
| 48 | Ivars Silamikelis, Kaspars Megnis, Monta Ustinova, Nikita Zrelovs, Vita Rovite, Mikus Gavars, Dmitrijs Perminovs, Uga Dumpis, Janis Kloviņš | hCoV-19/Latvia/05/2020 | E. Gulbja Laboratorija Latvian Biomedical Research and Study Centre Ivars SilamiÄ·elis et al https:__www.gisaid.org NA NA 4/15/2020 |
| 49 | Rita Feghali | hCoV-19/Lebanon/S9_764/2020 | Rafik Hariri University Hospital Rafik Hariri University Hospital Rita Feghali et al https:__www.gisaid.org NA NA 5/21/2020 |

| | | | |
|---|---|---|---|
| 50 | David Nieuwenhuijse, Bas Oude Munnink, Reina Sikkema, Claudia Schapendonk, Irina Chestakova, Anne van der Linden, Mark Pronk, Pascal Lexmond, Corien Swaan, Manon Haverkate, Madelief Mollers, Mart Stein, Sandra Kengne Kamga Mobou, Jeroen van Kampen, Jolanda Voermans, Aura Timen, Corine GeurtsvanKessel, Annemiek van der Eijk, Richard Molenkamp, Marion Koopmans, on behalf of the Dutch national COVID-19 response team. | hCoV-19/Netherlands/NoordBrabant_17/2020 | Dutch COVID-19 response team Erasmus Medical Center David Nieuwenhuijse et al https:__www.gisaid.org Rapid SARS-CoV-2 whole genome sequencing for informed public health decision making in the Netherlands https:__dx.doi.org_10.1038_s41591-020-0997-y 3/12/2020 |
| 51 | Oluniyi P.E., Ajogbasile F.V., Kayode A., Oguzie J., Olawoye I., Uwanibe J., Olumade T., Folarin O.A., Ihekweazu C., Happi C.T. | hCoV-19/Nigeria/ON507-CV44/2020 | Nigeria Centre for Disease Control (NCDC) African Centre of Excellence for Genomics of Infectious Diseases (ACEGID), Redeemer's University, Ede, Osun State, Nigeria Oluniyi P.E. et al https:__www.gisaid.org New SARS-CoV-2 Genomes from Nigeria Reveals Dominance of Viruses with Spike Protein Mutation (D614G), and Additional Virus Lineages in Circulation https:__virological.org_t_new-sars-cov-2-genomes-from-nigeria-reveals-dominance-of-viruses-with-spike-protein-mutation-d614g-and-additional-virus-lineages-in-circulation_527 7/13/2020 |
| 52 | Dorota Kujawa, Aleksandra Herud, Dariusz Martynowski, Krzysztof Jakub Pawlik, Joanna Sikorska, Paulina Zebrowska, Grazyna Zalewska, Oskar Karpinski and Lukasz Laczmanski | hCoV-19/Poland/Wro-02/2020 | Laboratory of Genomics & Bioinformatics, Institute of Immunology and Experimental Therapy, Polish Academy of Sciences Oddzial Mikrobiologii Wojewodzkiej Stacji Sanitarno-Epidemiologicznej. Laboratory of Genomics & Bioinformatics, Institute of Immunology and Experimental Therapy, Polish Academy of Sciences Dorota Kujawa et al https:__www.gisaid.org NA NA 5/11/2020 |
| 53 | Guiomar et al | hCoV-19/Portugal/PT0037/2020 | H Braga Instituto Nacional de Saude (INSA) Guiomar et al et al https:__www.gisaid.org NA NA 3/28/2020 |
| 54 | Abdullatif Al-Khal, Muna A. S. Al-Maslamani, Ajaeb D. M. H. Al-Nabet, Peter V. Coyle, Einas A. E. Al-Kuwari, Nourah B. M. Younes, Hamad E. Al-Romaihi, Salih Al-Marri, Mohammed Al-Thani, Fatiha M. Benslimane, Heba A. Al-Khatib, Sonia Boughattas, Hadi M. Yassine, Asmaa A. Al-Thani. | hCoV-19/Qatar/QA16/2020 : hCoV-19/Qatar/QA22/2020 | Ministry of Public Health (MoPH) Biomedical Research Center (BRC) Abdullatif Al-Khal et al https:__www.gisaid.org NA NA 4/18/2020 |
| 55 | Alexey Shchetinin, Maria Nikiforova, Nadezhda Kuznetsova, Ekaterina Aksenova, Marina Kunda, Natalia Ryzhova, Olga Voronina, Inna Dolzhikova, Daria Grousova, Andrey Botikov, Denis Logunov, Alexander Gintsburg, Vladimir Gushchin | hCoV-19/Russia/Moscow_PMVL-1/2020 | Russian State Collection of Viruses Pathogenic Microorganisms Variability Laboratory Alexey Shchetinin et al https:__www.gisaid.org NA NA 4/7/2020 |
| 56 | A. Pavlenko, O. Guskova, K. Klimina, V. Veselovsky, A. Manolov, D. Fedorov, V. Govorun and E. Ilina | hCoV-19/Russia/Moscow-GCBL1/2020 | Genomics and Computational Biology Lab, Scientific Research Institute of Physical-Chemical Medicine, FMBA of Russia Genomics and Computational Biology Lab, Scientific Research Institute of Physical-Chemical Medicine, FMBA of Russia A. Pavlenko et al https:__www.gisaid.org NA NA 5/7/2020 |
| 57 | Andrey Komissarov, Artem Fadeev, Mariia Sergeeva, Anna Ivanova, Daria Danilenko | hCoV-19/Russia/StPetersburg-3524/2020 : hCoV-19/Russia/StPetersburg-RII3992/2020 : hCoV-19/Russia/StPetersburg-RII4382V/2020 : hCoV-19/Russia/StPetersburg-RII4532V/2020 : hCoV-19/Russia/StPetersburg- | WHO National Influenza Centre Russian Federation WHO National Influenza Centre Russian Federation Andrey Komissarov et al https:__www.gisaid.org NA NA 3/19/2020 |

| | | | |
|---|---|---|---|
| | | RII4546V/2020 : hCoV-19/Russia/StPetersburg-RII4988S/2020 : hCoV-19/Russia/StPetersburg-RII5169S/2020 : hCoV-19/Russia/StPetersburg-RII5244S/2020 : hCoV-19/Russia/StPetersburg-RII6053S/2020 : hCoV-19/Russia/Ulan-Ude-RII4562V/2020 | |
| 58 | Ana da Silva Filipe, Kathy Smollett, Stephen Carmichael, Natasha Johnson, Daniel Mair, Lily Tong, Jenna Nichols; Sarah McDonald; Richard Orton, Joseph Hughes, Sreenu Vattipally, David L Robertson; Kathy Li, Natasha Jesudason, Rajiv Shah, James Shepherd, Antonia Ho, Emma Thomson; Alasdair MacLean, Rory Gunson. | hCoV-19/Scotland/CVR205/2020 | West of Scotland Specialist Virology Centre, NHSGGC _ MRC-University of Glasgow Centre for Virus Research COVID-19 Genomics UK (COG-UK) Consortium Ana da Silva Filipe et al https:__www.gisaid.org Preliminary analysis of SARS-CoV-2 importation & establishment of UK transmission lineages https:__virological.org_t_preliminary-analysis-of-sars-cov-2-importation-establishment-of-uk-transmission-lineages_507 4/14/2020 |
| 59 | Shengyue Wang, Xiaonan Zhang, Gang Lu, Yun Tan, Yun Ling, Hongzhou Lu, Saijuan Chen | hCoV-19/Shanghai/SH0030/2020 | Shanghai Public Health Clinical Center, Shanghai Medical College, Fudan University National Research Center for Translational Medicine (Shanghai), Ruijin Hospital affiliated to Shanghai Jiao Tong University School of Medicine & Shanghai Public Health Clinical Center Shengyue Wang et al https:__www.gisaid.org NA NA 3/20/2020 |
| 60 | Mak TM, Octavia S, Chavatte JM, Cui L, Lin RTP | hCoV-19/Singapore/67/2020 | National Public Health Laboratory, National Centre for Infectious Diseases National Public Health Laboratory, National Centre for Infectious Diseases Mak TM et al https:__www.gisaid.org NA NA 4/22/2020 |
| 61 | Octavia S, Mak TM, Cui L, Lin RTP | hCoV-19/Singapore/10/2020 : hCoV-19/Singapore/9/2020 | National Public Health Laboratory, National Centre for Infectious Diseases National Centre for Infectious Diseases, National Centre for Infectious Diseases Octavia S et al https:__www.gisaid.org NA NA 2/18/2020 |
| 62 | Monika Slávikova, Martina Ličková, Sabina Fumačová Havlíková, Juraj Koči, Juraj Kopáček, Elena Tichá, Edita Staroňová, Jaroslav Budiš, Werner Krampl, Miroslav Böhmer, Diana Rusňáková, Tomáš Szemeš, Boris Klempa | hCoV-19/Slovakia/SK-BMC2/2020 | Institute of Virology, Biomedical Research Center of the Slovak Academy of Sciences, Bratislava; Public Health Authority of the Slovak Republic, Bratislava Institute of Virology, Biomedical Research Center of the Slovak Academy of Sciences, Bratislava; Comenius University Science Park, Bratislava Monika SlÃ¡vikova et al https:__www.gisaid.org NA NA 3/27/2020 |
| 63 | Changmin Kang, Joon-Yong Bae, Jungmin Lee, Heedo Park, Juyoung Cho, Jeonghun Kim, Gee eun Lee, Cui Chunguang, Kyeong-ryeol Shin, Dong Min Kim, Jin Il Kim, Man-Seong Park | hCoV-19/South Korea/KUMC01/2020 : hCoV-19/South Korea/KUMC02/2020 | Department of Microbiology, Institute for Viral Diseases, College of Medicine, Korea University Department of Microbiology, Institute for Viral Diseases, College of Medicine, Korea University Changmin Kang et al https:__www.gisaid.org NA NA 3/3/2020 |
| 64 | Park,W.B., Kwon,N.-J., Choi,S.-J., Kang,C.K., Choe,P.G., Kim,J.Y., Yun,J., Lee,G.-W., Seong,M.-W., Kim,N., Seo,J.-S. and Oh,M.-D. | hCoV-19/South Korea/SNU01/2020 | unknown Department of Clinical Diagnostics Park et al https:__www.gisaid.org Virus Isolation from the First Patient with SARS-CoV-2 in Korea https:__dx.doi.org_10.3346_jkms.2020.35.e84 2/21/2020 |
| 65 | Iglesias-Caballero, M. Molinero Calamita, M. González-Esguevillas, M. Camarero, S. Pozo, F. Casas, I. Jiménez, P. Jiménez, M. Zaballos, A. Monzón, S. Varona, S. Juliá, M. Cuesta, I. Sanbonmatsu S. | hCoV-19/Spain/AN-ISCIII-201373/2020 : hCoV-19/Spain/AN-ISCIII-201649/2020 | HOSPITAL UNIVERSITARIO VIRGEN DE LAS NIEVES Instituto de Salud Carlos III Iglesias-Caballero et al https:__www.gisaid.org NA NA 3/29/2020 |

| | | | |
|---|---|---|---|
| 66 | Iglesias-Caballero, M. Molinero Calamita, M. González-Esguevillas, M. Camarero, S. Pozo, F. Casas, I. Jiménez, P. Jiménez, M. Zaballos, A. Monzón, S. Varona, S. Juliá, M. Cuesta, I. Marcos M.A | hCoV-19/Spain/CT-ISCIII-201396/2020 | HOSPITAL CLINIC Instituto de Salud Carlos III Iglesias-Caballero et al https:__www.gisaid.org NA NA 3/29/2020 |
| 67 | Silvia Hernáez Crespo, Carmen Gómez González, Amaia Aguirre Quiñonero, Marina Fernández Torres, Mª Rosario Almela Ferrer, Mª Concepción Lecaroz Agara, Andrés Canut Blasco. and SeqCOVID-SPAIN consortium | hCoV-19/Spain/PV-IBV-001436/2020 | Hospital Universitario Araba. Vitoria-Gasteiz SeqCOVID-SPAIN consortium_IBV(CSIC) Silvia HernÃ¡ez Crespo et al https:__www.gisaid.org NA NA 6/12/2020 |
| 68 | Elias Dahdouh, Sara González, Raúl Recio, Fernando Lázaro, Esther Viedma, Natalia Stella, Julio García, Juan Carlos Galán, Rafael Cantón, Mª Dolores Folgueira, Rafael Delgado, Jesús Mingorance | hCoV-19/Spain/MD-HLP-LP15-4/2020 : hCoV-19/Spain/MD-H12-LP33/2020 | Hospital Universitario La Paz Hospital Universitario 12 de Octubre Elias Dahdouh et al https:__www.gisaid.org NA NA 4/27/2020 |
| 69 | David Navarro, Maria Alma Bracho, Giuseppe D'Auria, Griselda De Marco, Neris Garcia-Gonzalez, Fernando Gonzalez-Candelas | hCoV-19/Spain/VC-FISABIO-3/2020 | Servicio Microbiologia, Hospital Clinico Universitario, Valencia Sequencing and Bioinformatics Service and Molecular Epidemiology Research Group. FISABIO-Public Health. David Navarro et al https:__www.gisaid.org Phylodynamics of SARS-CoV-2 transmission in Spain https:__dx.doi.org_10.1101_2020.04.20.050039 3/14/2020 |
| 70 | Paula Ruiz-Hueso, Mariana Reyes-Prieto, Vicente Soriano Chirona, Maria Alma Bracho, Griselda De Marco, Beatriz Beamud, Lidia Ruiz Roldan, Marta Pla Diaz,Neris Garcia-Gonzalez, Loreto Ferrús Abad, Inma Galán Vendrell, Maria Dolores Ocete, Llúcia Martínez-Priego, Concepcion Gimeno, Giuseppe D'Auria, Fernando Gonzalez-Candelas | hCoV-19/Spain/VC-FISABIO-34/2020 | Servicio de MicrobiologÃa. Consorcio Hospital General Universitario de Valencia Sequencing and Bioinformatics Service and Molecular Epidemiology Research Group. FISABIO-Public Health Paula Ruiz-Hueso et al https:__www.gisaid.org NA NA 4/5/2020 |
| 71 | Eva Espmark, Oskar Karlsson Lindsjo, Maria Lind Karlberg, Anna-Malin Linde, Olov Svartstrom, Anna Risberg, Theresa Enkirch, Mia Brytting, Karin Tegmark-Wisell | hCoV-19/Sweden/20-06308/2020 | Ektorps Vardcentral The Public Health Agency of Sweden Eva Espmark et al https:__www.gisaid.org NA NA 4/30/2020 |
| 72 | Anna-Malin Linde, Maria Lind Karlberg, Mattias Haukland, Reza Advani, Olov Svartstrom, Oskar Karlsson Lindsjo, Petra Edquist, Shamam Muradrasoli, Anna Risberg, Karin Tegmark-Wisell | hCoV-19/Sweden/20-08040/2020 | Halmstad klinisk mikrobiologi The Public Health Agency of Sweden Anna-Malin Linde et al https:__www.gisaid.org NA NA 5/28/2020 |
| 73 | Christian Beisel, Sarah Nadeau, Ivan Topolsky, Pedro Ferreira, Philipp Jablonski, Susana Posada-Céspedes, Tobias Schär, Ina Nissen, Natascha Santacroce, Elodie Burcklen, Christiane Beckmann, Maurice Redondo, Olivier Kobel, Christoph Noppen, Sophie Seidel, Noemie Santamaria de Souza, Niko Beerenwinkel, Tanja Stadler | hCoV-19/Switzerland/ZH-ETHZ-130026/2020 | Viollier AG Department of Biosystems Science and Engineering, ETH ZÃ¼rich Christian Beisel et al https:__www.gisaid.org NA NA 6/12/2020 |
| 74 | Ahmad Abou Tayoun, Tom Loney, Hamda Khansaheb, Sathishkumar Ramaswamy, Divinlal Harilal, Zulfa Omar Deesi, Rupa Murthy Varghese, Hanan Al Suwaidi, Abdulmajeed Alkhaja, Mohammed Uddin, Rifat Hamoudi, Rabih Halwani, Abiola Catherine Senok, Qutayba Hamid, Norbert Nowotny, Alawi Alsheikh-Ali | hCoV-19/United Arab Emirates/L0/2020 : hCoV-19/United Arab Emirates/L0184/2020 : hCoV-19/United Arab Emirates/L068/2020 : hCoV-19/United Arab Emirates/L0881/2020 : hCoV-19/United Arab Emirates/L0904/2020 : hCoV-19/United Arab Emirates/L2185/ | Mohammed Bin Rashid University of Medicine and Health Sciences Al Jalila Genomics Center Ahmad Abou Tayoun et al https:__www.gisaid.org Genomic surveillance and phylogenetic analysis reveal multiple introductions of SARS-CoV-2 into a global travel hub in the Middle East https:__dx.doi.org_10.1101_2020.05.06.080606 5/3/2020 |

| | | | |
|---|---|---|---|
| | | 2020 : hCoV-19/United Arab Emirates/L2409/2020 : hCoV-19/United Arab Emirates/L3779/2020 : hCoV-19/United Arab Emirates/L4280/2020 : hCoV-19/United Arab Emirates/L5621/2020 : hCoV-19/United Arab Emirates/L5630/2020 : hCoV-19/United Arab Emirates/L9768/2020 | |
| 75 | Bin Fang, Xiang Li, Xiao Yu, Linlin Liu, Bo Yang, Faxian Zhan, Guojun Ye, Xixiang Huo, Junqiang Xu, Bo Yu, Kun Cai, Jing Li, Yongzhong Jiang. | hCoV-19/Wuhan/HBCDC-HB-02/2020 : hCoV-19/Wuhan/HBCDC-HB-03/2020 : hCoV-19/Wuhan/HBCDC-HB-04/2020 : hCoV-19/Wuhan/HBCDC-HB-05/2020 : hCoV-19/Wuhan/HBCDC-HB-06/2020 | The Central Hospital Of Wuhan Hubei Provincial Center for Disease Control and Prevention Bin Fang et al https:__www.gisaid.org Genome-wide data inferring the evolution and population demography of the novel pneumonia coronavirus (SARS-CoV-2) https:__dx.doi.org_10.1101_2020.03.04.976662 3/2/2020 : Union Hospital of Tongji Medical College, Huazhong University of Science and Technology Hubei Provincial Center for Disease Control and Prevention Bin Fang et al https:__www.gisaid.org Genome-wide data inferring the evolution and population demography of the novel pneumonia coronavirus (SARS-CoV-2) https:__dx.doi.org_10.1101_2020.03.04.976662 3/2/2020 : CR&WISCO GENERAL HOSPITAL Hubei Provincial Center for Disease Control and Prevention Bin Fang et al https:__www.gisaid.org A doubt of multiple introduction of SARS-CoV-2 in Italy: A preliminary overview https:__dx.doi.org_10.1002_jmv.25773 3/2/2020 : Wuhan Lung Hospital Hubei Provincial Center for Disease Control and Prevention Bin Fang et al https:__www.gisaid.org Genome-wide data inferring the evolution and population demography of the novel pneumonia coronavirus (SARS-CoV-2) https:__dx.doi.org_10.1101_2020.03.04.976662 3/2/2020 |
| 76 | Zhang,Y.-Z., Wu,F., Chen,Y.-M., Pei,Y.-Y., Xu,L., Wang,W., Zhao,S., Yu,B., Hu,Y., Tao,Z.-W., Song,Z.-G., Tian,J.-H., Zhang,Y.-L., Liu,Y., Zheng,J.-J., Dai,F.-H., Wang,Q.-M., She,J.-L. and Zhu,T.-Y. | hCoV-19/Wuhan/Hu-1/2019 | unknown National Institute for Communicable Disease Control and Prevention (ICDC) Chinese Center for Disease Control and Prevention (China CDC) Zhang et al https:__www.gisaid.org A new coronavirus associated with human respiratory disease in China https:__dx.doi.org_10.1038_s41586-020-2008-3 1/12/2020 |
| 77 | Lili Ren, Jianwei Wang, Qi Jin, Zichun Xiang, Zhiqiang Wu, Chao Wu, Yiwei Liu | hCoV-19/Wuhan/IPBCAMS-WH-01/2019 | Institute of Pathogen Biology, Chinese Academy of Medical Sciences & Peking Union Medical College Institute of Pathogen Biology, Chinese Academy of Medical Sciences & Peking Union Medical College Lili Ren et al https:__www.gisaid.org NA NA 1/11/2020 |
| 78 | Weijun Chen, Yuhai Bi, Weifeng Shi and Zhenhong Hu | hCoV-19/Wuhan/WH01/2019 : hCoV-19/Wuhan/WH04/2020 | General Hospital of Central Theater Command of People's Liberation Army of China BGI & Institute of Microbiology, Chinese Academy of Sciences & Shandong First Medical University & Shandong Academy of Medical Sciences & General Hospital of Central Theater Command of People's Liberation Army of China Weijun Chen et al https:__www.gisaid.org Genomic characterisation and epidemiology of 2019 |
| 79 | Si,H., Zhu,Y., Lin,H., Xie,S., Shi,Z., Zhou,P. | hCoV-19/Wuhan/OS52/2020 : hCoV-19/Wuhan/YB012602/2020 | unknown CAS Key Laboratory of Special Pathogens and Biosafety and Center for Emerging Infectious Diseases Si et al https:__www.gisaid.org NA NA 3/30/2020 |

**Supplementary References**

1 Kalendar R, Khassenov B, Ramankulov Y, Samuilova O, Ivanov KI. FastPCR: An in silico tool for fast primer and probe design and advanced sequence analysis. *Genomics* 2017; **109**: 312–9.

2 Babraham Bioinformatics - FastQC A Quality Control tool for High Throughput Sequence Data. http://www.bioinformatics.babraham.ac.uk/projects/fastqc/ (accessed Dec 16, 2020).

3 Ewels P, Magnusson M, Lundin S, Käller M. MultiQC: summarize analysis results for multiple tools and samples in a single report. *Bioinformatics* 2016; **32**: 3047–8.

4 SeqKit: A Cross-Platform and Ultrafast Toolkit for FASTA/Q File Manipulation. https://journals.plos.org/plosone/article?id=10.1371/journal.pone.0163962 (accessed Dec 16, 2020).

5 Li H. Aligning sequence reads, clone sequences and assembly contigs with BWA-MEM. *arXiv:13033997 [q-bio]* 2013; published online May 26. http://arxiv.org/abs/1303.3997 (accessed Dec 16, 2020).

6 Li H, Handsaker B, Wysoker A, *et al.* The Sequence Alignment/Map format and SAMtools. *Bioinformatics* 2009; **25**: 2078–9.

7 bcftools. http://www.htslib.org/doc/bcftools.html (accessed Dec 16, 2020).
